# Spatiotemporal variation in risk of *Shigella* infection in childhood: a global risk mapping and prediction model using individual participant data

**DOI:** 10.1101/2022.08.04.22277641

**Authors:** Hamada S. Badr, Josh M. Colston, Nhat-Lan H. Nguyen, Yen Ting Chen, Syed Asad Ali, Ajit Rayamajhi, Syed M. Satter, Nguyen Van Trang, Daniel Eibach, Ralf Krumkamp, Jürgen May, Ayola Akim Adegnika, Gédéon Prince Manouana, Peter Gottfried Kremsner, Roma Chilengi, Luiza Hatyoka, Amanda K. Debes, Jerome Ateudjieu, Abu S. G. Faruque, M. Jahangir Hossain, Suman Kanungo, Karen L. Kotloff, Inácio Mandomando, M. Imran Nisar, Richard Omore, Samba O. Sow, Anita K. M. Zaidi, Nathalie Lambrecht, Bright Adu, Nicola Page, James A. Platts-Mills, Cesar Mavacala Freitas, Tuula Pelkonen, Per Ashorn, Kenneth Maleta, Tahmeed Ahmed, Pascal Bessong, Zulfiqar A. Bhutta, Carl Mason, Estomih Mduma, Maribel P. Olortegui, Pablo Peñataro Yori, Aldo A. M. Lima, Gagandeep Kang, Jean Humphrey, Robert Ntozini, Andrew J. Prendergast, Kazuhisa Okada, Warawan Wongboot, Nina Langeland, Sabrina J. Moyo, James Gaensbauer, Mario Melgar, Matthew Freeman, Anna N. Chard, Vonethalom Thongpaseuth, Eric Houpt, Benjamin F. Zaitchik, Margaret N. Kosek

**Author notes:** These authors contributed equally (Hamada S. Badr, Josh M. Colston). **Corresponding author:** Correspondence about this manuscript can be addressed to: Dr. Benjamin F. Zaitchik, Department of Earth and Planetary Sciences, Johns Hopkins Krieger School of Arts and Sciences, Baltimore, 21218, MA, USA.

## Abstract

**Background:** Diarrheal disease remains a leading cause of childhood illness and mortality and *Shigella* is a major etiological contributor for which a vaccine may soon be available. This study aimed to model the spatiotemporal variation in pediatric *Shigella* infection and map its predicted prevalence across low- and middle-income countries (LMICs).

**Methods:** Independent participant data on *Shigella* positivity in stool samples collected from children aged ≤59 months were sourced from multiple LMIC-based studies. Covariates included household- and subject-level factors ascertained by study investigators and environmental and hydrometeorological variables extracted from various data products at georeferenced child locations. Multivariate models were fitted, and prevalence predictions obtained by syndrome and age stratum.

**Findings:** 20 studies from 23 countries contributed 66,563 sample results. Age, symptom status, and study design contributed most to model performance followed by temperature, wind speed, relative humidity, and soil moisture. *Shigella* probability exceeded 20% when both precipitation and soil moisture were above average and had a 43% peak in uncomplicated diarrhea cases at 33°C temperatures, above which it decreased. Improved sanitation and open defecation decreased *Shigella* odds by 19% and 18% respectively compared to unimproved sanitation.

**Interpretation:** The distribution of *Shigella* is more sensitive to climatological factors like temperature than previously recognized. Conditions in much of sub-Saharan Africa are particularly propitious for *Shigella* transmission, though hotspots also occur in South and Central America, the Ganges–Brahmaputra Delta, and New Guinea. These findings can inform prioritization of populations for future vaccine trials and campaigns.

**Funding:** NASA 16-GEO16-0047; NIH-NIAID 1R03AI151564-01; BMGF OPP1066146.

## Introduction

Diarrheal diseases remain a leading cause of childhood illness responsible for some 573,000 deaths in children under 5 in 2019 and caused by numerous infectious pathogens including bacterial, viral, and parasitic organisms.^1^ Moreover, many such enteropathogens have large reservoirs of asymptomatically infected individuals, and even in the absence of clinical diarrheal symptoms, repeated subclinical episodes can still impair growth^2–4^, cognitive development^5^ and vaccine response^6^ and may predispose children to chronic disease later in life.^7^ *Shigella*, a genus of gram-negative bacteria with 50 serotypes^8^, infects over 163 million people annually in low- and middle-income countries (LMICs)^9^ and is responsible for over 212,000 diarrheal disease deaths.^10^ It is most commonly recognized via its manifestation as bacillary dysentery, with 64,000^11^ such deaths occurring in under 5s, and an estimated case fatality of 2 per 1,000 in that age group.^12^ *Shigella* transmission is directly associated with temperature and is common in rural areas and where environmental risk factors such as poor water quality, lack of access to care, and inadequate sanitation are prevalent.^11,13–16^ Vaccine development for *Shigella* has been hampered by biotechnical and financial limitations, but there are multiple candidate vaccines now in late stages of development.^17^ It is therefore becoming increasingly important to geographically map global *Shigella* infection risk to guide and inform prospective rollout efforts towards high-priority populations.^18–21^

Recent years have seen burgeoning in the field of infectious disease cartography facilitated by advances in computing, remote sensing, geostatistical methods and the availability of spatially referenced environmental data.^10^ High-resolution model-based maps with continent-wide or quasi-global coverage have been generated, quantifying the spatial distribution of infectious pathogens including *Plasmodium falciparum*^23^ and *P. vivax*^24^, lymphatic filariasis^25^, and leishmaniases^26^ as well as all-cause diarrhea.^27^ However, attempts to map *Shigella* have hitherto been limited to indirect estimation methods. One study used estimates of overall diarrhea-related mortality and morbidity and adjusted these by published pathogen-specific attributable fractions of diarrhea with *Shigella* etiology to arrive at national-level incidence and death rates.^11^ Another used a similar approach, but focused on 11 African countries to derive subnational, province-level estimates based partly on covariates ascertained through household surveys.^28^ Notably, these analyses did not attempt to account for intra-annual seasonal and temporal variation in transmission, or spatial variation below the administrative unit level. Also, in focusing on morbidity and mortality, they could not quantify the contribution of asymptomatic infections that act as reservoirs for transmission.

This study aims for the first time to: 1) model the spatiotemporal variation in childhood *Shigella* infection probability using covariates with quasi-global coverage and gain insights into pathogen transmission, risk, and seasonality: 2) use the resulting estimates to map the predicted prevalence of the pathogen sub-nationally across LMICs.

## Methods

### Objective and scope

The objective of this analysis was to estimate the percent prevalence of enteric *Shigella* infection in three separate age groups 0-11, 12-23, and 24-59 months - and three syndrome strata – asymptomatic, uncomplicated (community detected) diarrhea, and medically attended diarrhea – at all locations throughout the world’s LMICs (as defined in the **supplementary appendix**).

### Data sources and outcome variable

To compile a dataset representative of diverse geographical and climatic contexts, data were sourced and compiled from multiple completed studies identified through non-systematic, exploratory literature review and professional networks according to inclusion criteria and within an Independent Participant Data Meta-Analyses (IPD-MA) framework described previously and in the **supplementary appendix**.^29^ 20 studies contributed data from 23 countries with a range of latitudes spanning the tropics and sub-tropics, including locations in Central and South America, sub-Saharan Africa and South and Southeast Asia (**figure 1**). The outcome of interest was *Shigella* infection status ascertained by polymerase chain reaction (PCR) performed on diarrheal and surveillance (asymptomatic) stool samples collected from children aged ≤59 months and was treated as a binary variable (positive or negative). The *Shigella* stool positivity rate (the probability of PCR-detection) was modeled as an approximation of the prevalence of pediatric *Shigella* infection. Discrete infection episodes were defined as previously described.^29^ **Figure 1** shows the locations, number of samples and the proportion positive for *Shigella* at each study site, while **supplementary table S1** summarizes key features of contributing studies.

**Figure 1.**
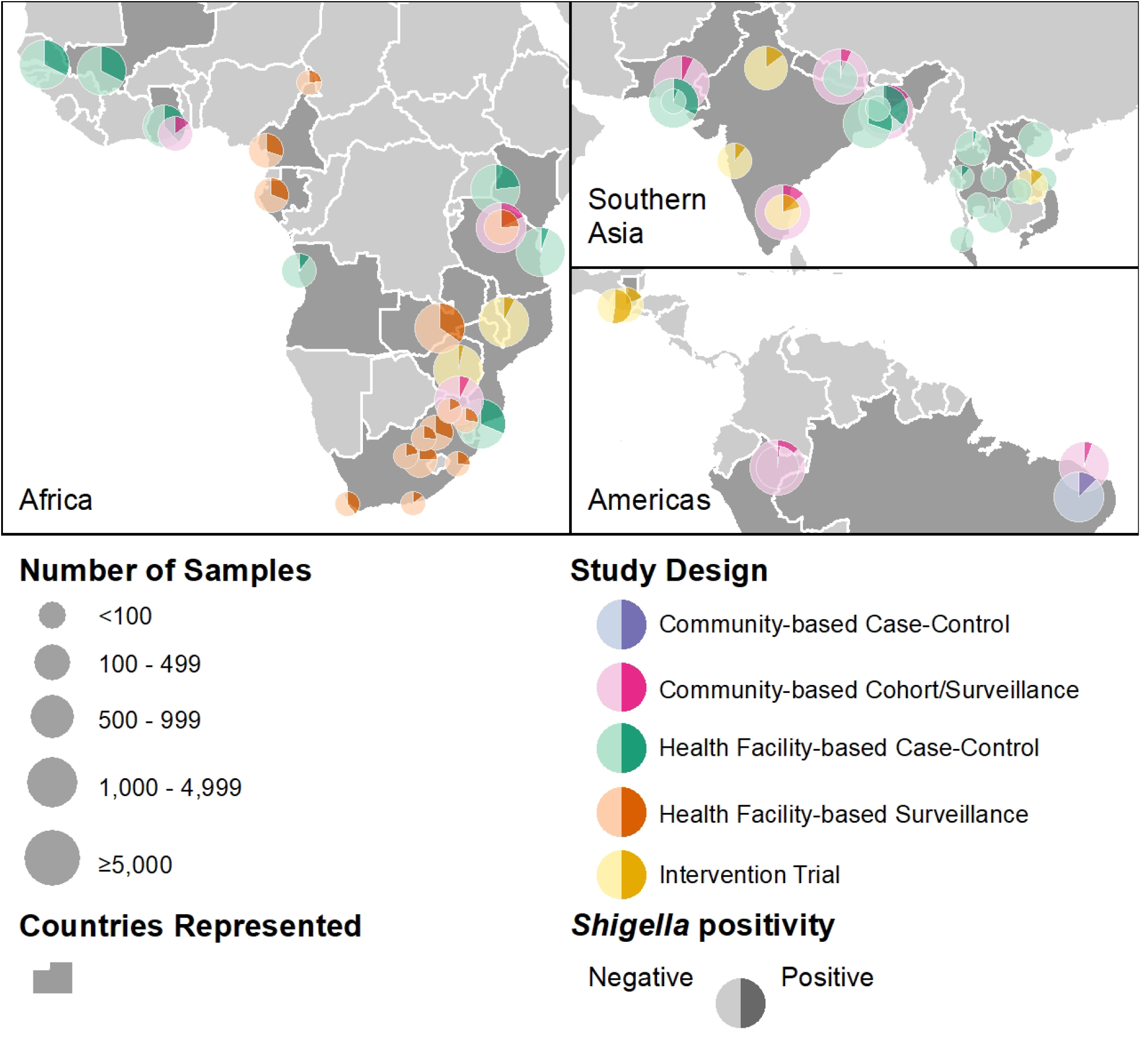
Locations of the sites and designs of the studies contributing data to this analysis, number of samples from each site included in the analysis and *Shigella*-positivity rates.

### Covariates

#### Symptom status and study design

13 (65%) of the included studies had health facility-based case-control or surveillance designs which actively recruited cases of gastrointestinal syndromes ranging in severity from uncomplicated to acute, often watery diarrhea and dysentery (bloody diarrhea). Results for diarrheal samples from symptomatic individuals were therefore overrepresented in the overall database relative to the frequency of diarrhea occurrence in the general population, which could lead to overestimation of risk for *Shigella*, a pathogen strongly associated with diarrhea and dysentery.^30^ To adjust for this, a categorical variable was included indicating whether the stool sample was collected while the child was asymptomatic or symptomatic - experiencing a diarrheal episode of any severity - and whether the sample was from a study with a community surveillance- or health facility-based design (four categories, representing each combination of the two symptom statuses and two study designs). This was to account for the assumed differential pathogen positivity rates in symptomatic samples and, especially, diarrhea for which facility care was sought.^29^ The inclusion of these terms allowed the predicted stool *Shigella*-positivity rate to be modeled separately for three symptom status categories: 1) asymptomatic; 2) community detected diarrhea; and 3) medically attended diarrhea (cases identified in health facility patients).

#### Age

Subject ages at sample collection were categorized into three age groups (0-11, 12-23, and 24-59 months) to adjust for the well-documented age-dependence of childhood *Shigella* risk and predict risk separately for age-groups commonly reported in studies of enteropathogen burden.^31^ Age, symptom status, and study design are hereafter referred to collectively as the “control variables”, since they were included in the model to control for confounding and are the only variables for which separate predictions were made for each value, rather than letting their values vary spatially.

#### Subject- and household-level covariates

Most contributing studies conducted baseline and/or follow-up assessments of information relevant to *Shigella* transmission risk and vulnerability. These included children’s feeding^13,32^ and nutritional^33^ status— exclusively breastfed, fully weaned, stunting, underweight and wasting—and household characteristics^16^—flooring material, household crowding, caregiver education, sanitation facility, open defecation and water source. These data were recoded to match as closely as possible to standardly used definitions of variables (**supplementary table S2**), and, where missing or not collected by some studies, imputed or interpolated based on household survey data according to methods described previously and in the **supplementary appendix**.^16^

#### Environmental spatial covariates

A suite of time-static environmental and sociodemographic spatial covariates (summarized in **supplementary table S2**) available in raster format were compiled based on their hypothesized or demonstrated associations with diarrheal disease outcomes.^27^ These included accessibility to cities, cropland areas, distance to major river, elevation, enhanced vegetation index, growing season length, human footprint index, irrigated areas, population density and urbanicity. Variable values were extracted at each child’s georeferenced location according to methods described in the **supplementary appendix**. All continuous spatial covariates were normalized and standardized to their overall distributions.

#### Time-varying hydrometeorological variables

A set of historical daily Earth Observation- and model-based re-analysis-derived estimates of hydrometeorological variables (**supplementary table S2**) were selected based on their demonstrated or hypothesized potential to influence enteric pathogen transmission, extracted from version 2.1 of the Global Land Data Assimilation System (GLDAS)^34^, where appropriate, standardized to local distributions and summarized over a lagged period of exposure, using methods described previously and in the **supplementary appendix**.^29^ These included precipitation, relative and specific humidity, soil moisture, solar radiation, surface pressure and runoff, temperature and wind speed.

### Statistical analysis

#### Model fitting

Generalized multivariate models were fitted to the binary infection status outcome to estimate predictive probabilities of *Shigella* positivity using the *additive*^35^ and *bayesian*^36^ R packages (developed for this analysis and extended as general-purpose software tools). An initial generalized linear model (GLM) including only the control variables was fitted to the outcome to serve as a reference for intercomparisons with subsequent models of increasing complexity in a repeated *k*-fold cross-validation analysis, namely: a GLM which included all non-hydrometeorological variables modeled as linear; a GLM which included all variables modeled as linear; and a final generalized additive model (GAM) with splines specified for the hydrometeorological variables to allow for the previously documented non-linearity of their relationships with *Shigella*.^29^ The performance of the models was compared based on numerous in-sample and out-of-sample classification metrics (**figure 2a**.) using cross validation, and the importance of variables was identified for all categories (**figure 2b**.) using accumulated local effects (ALE)^37^ and the average expected marginal contribution (Shapley values).^38^ An interaction between air temperature and symptom status was specified, based on a hypothesis and evidence from exploratory analyses, that temperature differentially affects the probability of diarrheal samples being *Shigella*-positive compared to samples from asymptomatic individuals. Similarly, an interaction between precipitation and soil moisture was specified based on the hypothesis that associations with rainfall on diarrhea-causing agents may be subject to effect modification by antecedent wetness conditions, such that the impact of extreme rainfall is increased following drier conditions.^39^

**Figure 2.**
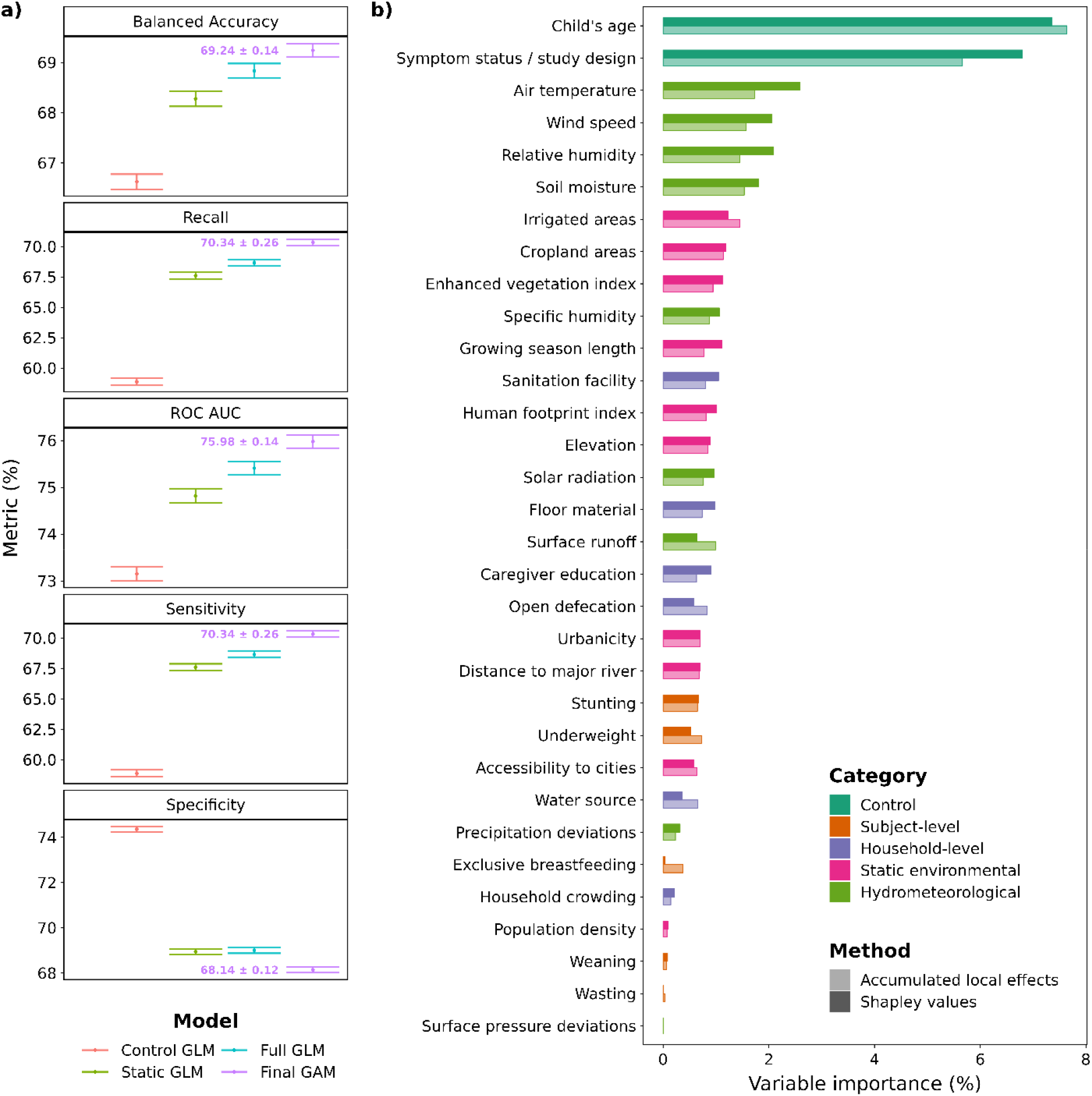
Variable importance plot for variables included in the full and final models

#### Model predictions

The final model results were used to make separate predictions for each syndrome and age stratum using the coefficient estimates for the corresponding control variables at each combination of their values. Raster files of the values of each other covariate across the geographic extent of the domain of interest, LMICs, were obtained or generated as described in the **supplementary appendix**. For the binary household- and subject-level covariates, coefficient estimates of the probability of *Shigella* infection in the comparison compared to the reference category were adjusted by the geographically varying proportion coverage or prevalence stored in raster format. Seasonality was explored by extracting and plotting time series of daily predictions at six key illustrative locations – settlements in areas of high and low prevalence in LMICs in each of the three regions – Africa, Asia, and the Americas.

Analyses were carried out using Stata 16^40^, R 4.0.3^41^ and ArcMap 10.8^42^ and PRISMA-IPD^43^ and GATHER^44^ guidelines were followed (see **supplementary appendix tables 4 and 5**).

## Results

**Table 1** shows the distribution of stool samples in the pooled database with available *Shigella* spp. positivity status included in this analysis by age group and child’s symptom status. Almost three quarters of diagnostic results were from surveillance samples, collected from asymptomatic individuals, while of the remaining diarrheal samples, almost twice as many were collected from children presenting at health facilities than those recruited in communities. Roughly equal numbers of samples were collected from children in the first and the second years of life, while those from older children (≥2 years) made up under ten percent of the database.

**Table 1.** Number (and percent) of stool samples with available Shigella spp. positivity status used in this analysis by child’s age and diarrhea symptom status. (Community detected diarrhea – diarrhea cases identified in community-based surveillance. Medically attended diarrhea – diarrhea cases identified in health facility-based studies).

| Age group | Diarrhea symptom status |  |  | Total |
| --- | --- | --- | --- | --- |
|  | Asymptomatic | Diarrheal symptoms |  |  |
|  |  | Community detected | Medically attended |  |
| 0 - 11 months | 23,013 (34.6) | 3,190 (4.8) | 4,796 (7.2) | 30,999 (46.6) |
| 12 - 23 months | 22,753 (34.2) | 2,577 (3.9) | 3,945 (6.0) | 29,275 (44.0) |
| 24 - 59 months | 3,514 (5.3) | 328 (0.5) | 2,447 (3.7) | 6,289 (9.4) |
| Total | 49,280 (74.0) | 6,095 (9.2) | 11,188 (16.9) | 66,563 (100) |

Figure 2 summarizes performance statistics of the final model (**2a**.) and two metrics of importance for each included variable – Shapley values and ALE – which are ranked by the average of the two (**2b**.). By every metric, the performance of the models improved with increasing complexity, and the final GAM that modeled the hydrometeorological variables as non-linear showed improved performance over the GLMs which treated them as linear. Child age at the time of sample collection was the most important contribution by both metrics, followed by the other control variable, symptom status/study design. The next four most important were all time-varying hydrometeorological variables (temperature, wind speed, relative humidity, and soil moisture), followed by three static environmental variables (irrigated and cropland areas and enhanced vegetation index). The most important household-level variable was presence of an improved sanitation facility, which ranked 12^th^, while the most important subject-level variable, stunting, ranked far down the list at 22^nd^.

Figure 3. shows the probability of *Shigella* infection predicted by the conditional effects of the most important hydrometeorological variables (and their interactions) in the final model (*3c*. and *3d*.) and the odds ratios for the other variables (**3a**. and **3b**.). Children aged 24 – 59 months had around 5-fold increased odds of *Shigella* positivity (odds ratio [OR]=4.82, confidence interval [4.43-5.21]) compared to those aged <1 year, while the equivalent effect in the second year of life was a more than 3 times increased odds (OR=3.42 [3.24-3.61]). Temperature exhibited a non-linear, asymmetrical, inverse U-shaped association with the probability of *Shigella* infection that was most marked for symptomatic children. Probability increased steadily across the first three quartiles of the temperature distribution, peaking at a value of around 34°C with a 47% probability of *Shigella* detection for uncomplicated diarrhea cases and 28% for asymptomatic children, before decreasing above that threshold. An interaction was observed between precipitation and soil moisture, such that probability of positivity was low across the entire range of the precipitation distribution during very dry soil conditions, and moderate during periods of below-average rainfall, but when both variables were above average concurrently, *Shigella* detection probability increased and exceeded 20%. Wind speed also had an asymmetrical, inverse U-shaped association with probability of a child having *Shigella* but skewed to the other side, with the peak risk occurring at speeds of 4m/s. Relative humidity and solar radiation had a broadly direct effect on *Shigella* risk, and specific humidity a low magnitude inverse association with the probability of *Shigella* detection.

Living in peri-urban areas had the largest direct association with *Shigella* of the non-hydrometeorological variables, a statistically significant 18% increased odds (OR=1.18 [1.07-1.29]) compared to rural residents, though the equivalent estimate for fully urban areas was non-significant. Two anthropometric markers, were next with moderate-to-severely underweight or stunted children having a statistically significant approximate 17% (OR=1.17 [1.08-1.26]) and 16% (OR=1.16 [1.09-1.23]) increased odds of testing positive for *Shigella* respectively. The other subject-level variables, exclusive breastfeeding, full weaning and wasting had negligible, non-significant effects. The largest inverse associations among the non-hydrometeorological variables were for five of the household-level covariates. Living in a household that has an improved sanitation facility or practices open defecation both decreased the odds of *Shigella* detection by just under 20% (respectively OR=0.81 [0.76-0.86], and OR=0.82 [0.76-0.88]) compared to an unimproved facility, while having improved floor material (OR=0.83 [0.78-0.88]), a caregiver who completed primary education (OR=0.85 [0.81-0.89]), or an improved water source (OR=0.86 [0.79-0.93]) had slightly smaller protective effects. Of the remaining environmental variables, irrigated areas (OR=1.13 [1.08-1.18]), growing season length (OR=1.11 [1.05-1.17]), enhanced vegetation index (OR=1.11 [1.05-1.17]), and elevation (OR=1.08 [1.03-1.13]) were associated with a statistically significantly increased risk of *Shigella* infection whereas cropland areas (OR=0.90 [0.86-0.94]), the human footprint index (OR=0.91 [0.85-0.97]), and distance to a major river (OR=0.94 [0.90-0.98]), were associated with a lower risk.

**Figure 3.**
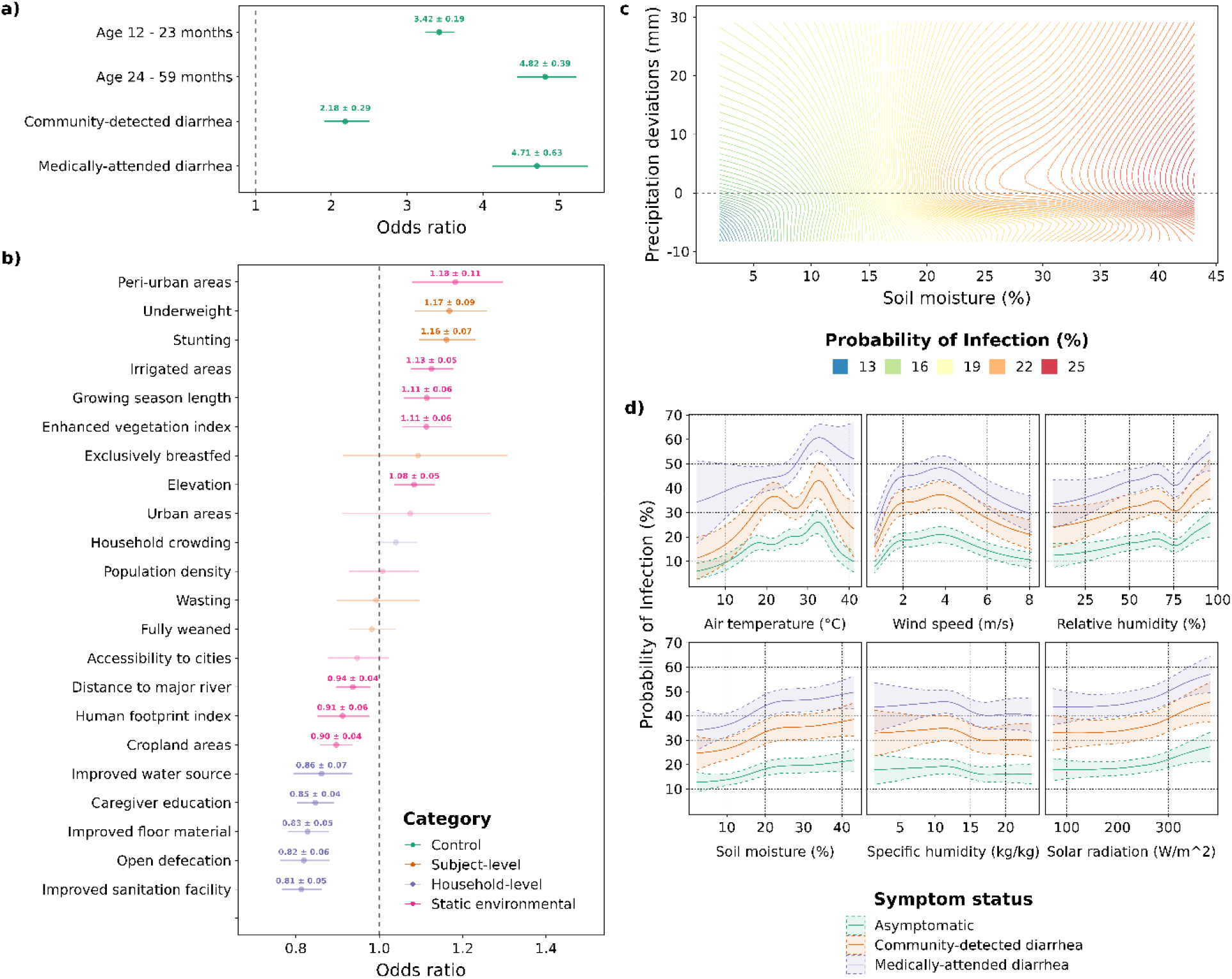
Probabilities and odds ratios of Shigella infection predicted by the conditional effects of each variable in the final model. The reference category for age is the 0 – 11- month group and the coefficients for the diarrhea categories are in comparison to asymptomatic children from community surveillance studies. Odds ratios for continuous static environmental variables are for one standard deviation increase. Precipitation deviations are relative to long term (2005-2019) site-specific averages.

Figure 4 shows the geographical distribution of the annual average prevalence of *Shigella* positivity predicted by the model for children aged 12 – 23 months in 2018 for each of the three symptom strata. Predicted prevalence of *Shigella* in asymptomatic individuals (**4a**.) varied from below 2.5% in areas of western China, Central Asia, and the Argentinian Andes to over 20% in small pockets of Central America (notably eastern Nicaragua), northern South America, tropical sub-Saharan Africa, Bangladesh, Southeast Asia (notably northeastern Cambodia), and Papua New Guinea. For individuals with uncomplicated diarrhea (**4b**.), the distribution was shifted upward such that there were large areas with predicted prevalence of higher than 25%, including almost all the territories of the Central African Republic, and South Sudan. Large areas of high prevalence were also predicted across the other countries around the African Great Lakes, coastal West Africa, Angola, Madagascar, and Bangladesh, with more delimited pockets discernable in Caribbean Central America, the interiors of Bolivia, Colombia and Venezuela and continental Southeast Asia. The predicted prevalence in individuals with medically attended diarrhea (**4c**.) exceeded 30% over almost the entirety of the tropics (with some exceptions such as parts of Borneo and the Andes) as well as large areas of subtropical Mexico, eastern China, and northeastern Argentina. Across all three symptom strata, the lowest risk was predicted over the greater Central Asia region (including Mongolia and western China), Saharan North Africa, the Andes, and Southern Africa.

**Supplementary animation S1** shows the temporal variation in the predictions for each day of the year 2018, while **supplementary figure S4** shows the annual time series of daily and smoothed predictions for asymptomatic children aged 12-23 months over the same year at six illustrative locations. Seasonal patterns varied. The high-prevalence Central African Republic location showed a boreal spring peak in early April, compared with the low-prevalence Botswana location where a December to February peak was evident. The high-prevalence Papua New Guinea location showed low amplitude, with only slightly higher values in the March-October period, while in Bishkek, Kyrgyzstan, a more marked mid-year peak of 11% was observed, subsiding to very low prevalence at the beginning and end of year. A mid-year and secondary end-of-year peak was discernable in high-prevalence Bluefields, Nicaragua, in contrast to La Paz, Bolivia where year-round low prevalence showed little seasonal variation. Seasonality metrics are mapped in **supplementary figure S5**.

**Figure 4.**
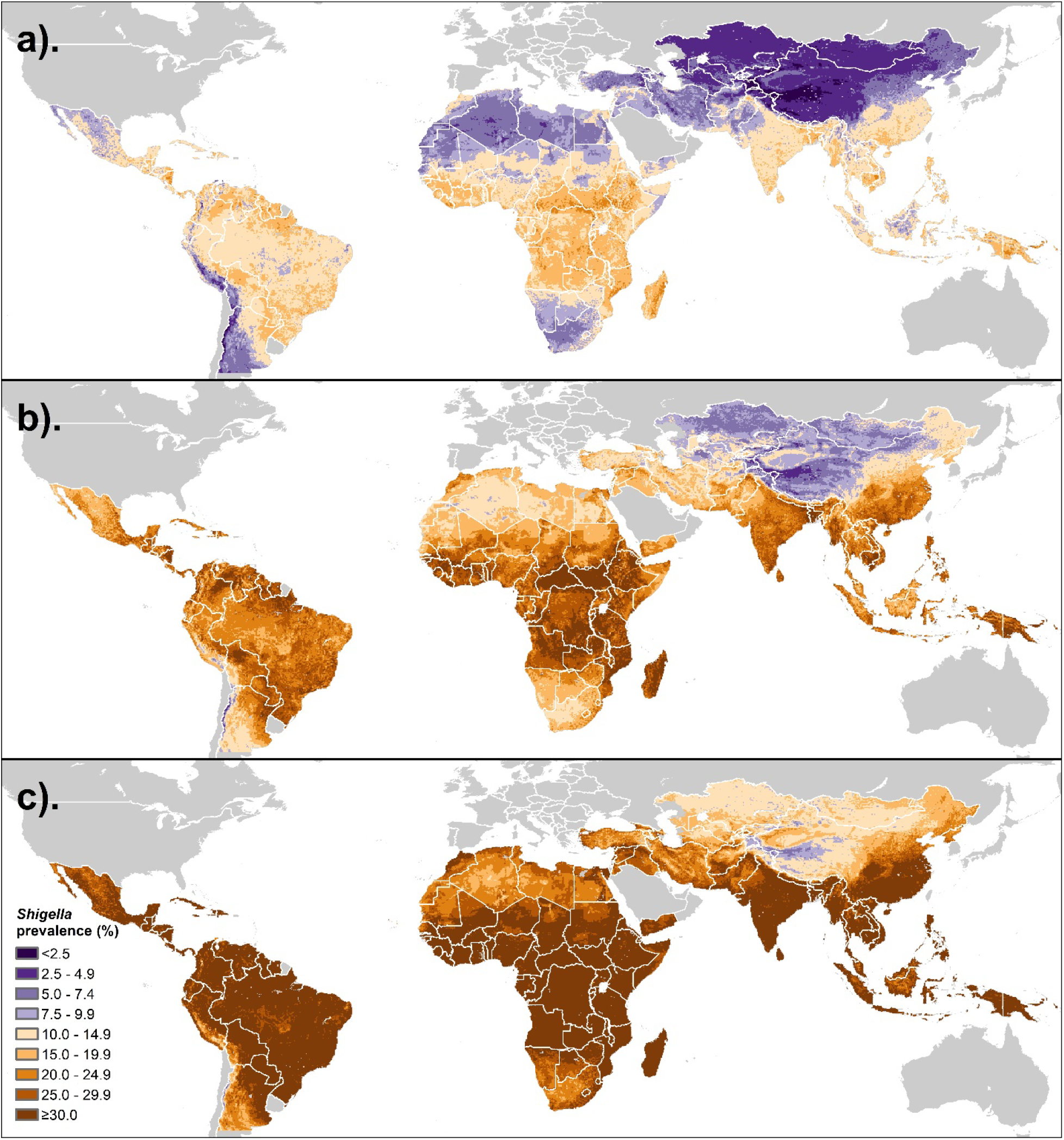
Geographical distribution of the annual average prevalence of *Shigella* infection in children aged 12 – 23 months predicted by the final model in a). Asymptomatic individuals; b). Community detected diarrhea cases; c). Medically attended diarrhea cases

## Discussion

Disease burden indicators for numerous high priority pathogens have been comprehensively mapped over entire endemic regions.^45–47^ However, pathogen-specific enteric infectious diseases have largely been excluded from such efforts^48,49^, despite calls for innovative approaches to such estimation to inform future vaccine and other programs.^19,50,51^ This is in part due to a perceived lack of readily accessible, spatially referenced data on their detection, since enteropathogen cases are not routinely reported through health information systems or household surveys.^52^ Although newly developed multiplex molecular diagnostic platforms are increasingly being deployed in surveillance studies to detect and differentiate an array of microbes in stool samples,^53–55^ studies of this nature have diverse designs and no single one offers a sufficiently broad range of geographical and environmental contexts. This study applied an IPD-MA approach to address this knowledge gap and is the first attempt to map *Shigella* prevalence at sub-unit level and across multiple continents. It is also the first to model the comparative effects of a large suite of covariates that vary on different scales, from the individual to the wider macroclimate, and, for several, include temporal variability at daily resolution. The data sources and analytic methods were selected to support spatial inference, resulting in a dataset of unparalleled scale and scope, and allowing for the first time to derive spatiotemporally complete, quasi-global predictions of *Shigella* infection rates as a function of climatic, environmental, and socio-demographic factors that can be extracted at specific locations. The findings reveal not only small-scale zones of potential elevated transmission risk, but also generalizable evidence about the relative influence of different drivers of *Shigella* transmission. Furthermore, the seasonal variability in infection risk within the same locations revealed by these findings may be relevant for diagnosis and treatment and for optimal timing of health system interventions.

The results confirm that *Shigella* risk has a strong association with age – the highest prevalence occurring in older children (24-59m) - and diarrhea symptom status, consistent with effects documented previously.^13,30,56^ While these control variables ranked highest in importance and effect size magnitude, they were closely followed by hydrometeorological variables including temperature, wind speed, relative humidity, and soil moisture. These associations exhibited considerable non-linearity and were consistent in shape, though larger in magnitude, than those identified from an earlier version of this dataset.^29^ Atmospheric and hydrological factors likely impact the survival and dispersal of bacteria like *Shigella* outside the host where warm and moist conditions may prolong their viability.^29,57^ The importance of temperature in *Shigella* transmission also confirms previously reported findings. A meta-analysis found a relative risk (RR) of *Shigella* diagnosis of 1.07 for each 1°C increase in temperature^58^, while the equivalent for a 5°C increase in data pooled from several hundred sentinel sites across China was 1.31.^59^ However, we also demonstrate that *Shigella* prevalence drops sharply in the upper temperature extreme above a value of around 33°C, and that the inverse U-shape of this association is more pronounced for symptomatic than asymptomatic *Shigella*.

While some studies have found no association between rainfall and *Shigella* infection or shigellosis^59,60^, here not only was a direct effect of precipitation observed, but also an interaction with soil moisture such that prevalence was highest when both parameters were above their average values. Kraay and colleagues hypothesize that extreme rainfall increases diarrhea incidence to a greater extent if it follows a drier period.^39^ These findings suggest that such an effect, if true, is not mediated by *Shigella*. This compounding of soil saturation conditions on precipitation is, however, consistent with findings from an interrupted time-series analysis of a large, La Niña-related flood in the study site in Loreto, Peru, where an increase in *Shigella* infection was observed, but only in the later period of the flood.^61^

While household factors ranked lower in importance, several statistically significant protective associations of these variables were observed. Living in a household that practices open defecation (i.e., has no sanitation facility) conferred lower odds of Shigella infection by almost one fifth (18%), a protective association almost equal to that of having an improved sanitation facility (19%). This runs counter to the widely held perception that open defecation promotes transmission of infectious intestinal diseases,^62^ and carries the implication that, from the perspective of childhood *Shigella* prevention, having no toilet at all is preferable to an inadequate one (though open defecation may still be a risk factor for other diarrheal pathogens and adverse health and social outcomes).^63^ These effects sizes are smaller than those from a trial of pour-flush toilets with septic tanks in Maputo, Mozambique, which reduced the prevalence of *Shigella* in children under 2 years by half.^64^ While the further protective effects of caregiver primary education and covered floors (both ∼15%) are consistent with those reported previously from an earlier version of this dataset^65^, the 14% reduced odds of *Shigella* infection conferred by having an improved water source is modest compared with findings from Manhiça District Mozambique that water availability is among the most protective factors against *Shigella* infection in under 2s.^66^

The direct association of two indicators of undernutrition, moderate-to-severe underweight and stunting, were of comparable magnitude to the protective effects of household factors, meaning that children with a weight- or height-for-age Z-score (WAZ and HAZ) 2 or more standard deviations below that of a healthy reference population were at increased risk of *Shigella* infection. This is further evidence of the well-documented bidirectional interplay between malnutrition and enteric infections^2,4,13,67^, and consistent with findings from multi-site studies that both symptomatic^33^ and asymptomatic^68^, untreated *Shigella* infections in young children are associated with later adverse anthropometric outcomes. It is notable that feeding status was not statistically significantly associated with *Shigella* positivity, since early introduction of complementary foods has been shown to be associated with a 10% increased RR of infection with *Shigella*.^13^ However, it is consistent with a comparative analysis of the effect of full breastfeeding on multiple enteropathogens which found associations with some viruses and bacteria, but a non-significant association with *Shigella*.^32^

This study had various limitations. It was only possible to directly estimate *Shigella* PCR-positivity in diarrheal samples and not the *Shigella*-specific attributable burden of diarrhea, which, due to the pervasiveness of coinfection with multiple diarrhea-causing agents, are far from equivalent, the former tending to overestimate the latter considerably. The prediction estimates for children with diarrhea provided in supplementary files can be adjusted downward using generalizable or context-specific *Shigella*-attributable fraction as these become available. Similarly, these prevalence estimates should not be conflated with incidence, which could be higher in settings where repeated diarrheal episodes are common even where point prevalence is low. Potential sources of error include the use of interpolation and imputation of missing subject- and household-level data due to inconsistency of covariate ascertainment across contributing studies, as well as the use of recruitment center coordinates as proxies for georeferencing children’s residence locations where the latter were unavailable (see **supplementary materials**). Furthermore, the relatively coarse resolution of the GLDAS estimates restricted the precision of the prevalence predictions to a horizontal resolution of around 25km^2^. However, the consistency of many major findings with those established in the prior literature give cause for confidence that any such error would be so minimal as to not bias the broad conclusions.

In conclusion, the spatiotemporal distribution of *Shigella* risk in LMICs appears highly sensitive to both macro-scale and inter-diurnal variation in temperature and other climatological factors. This makes conditions in sub-Saharan Africa particularly propitious for propagation of the bacterium. Bands of elevated prevalence were predicted to the immediate north of the Congo basin and Great Lakes region, just to the north of the Tropic of Capricorn, and in tropical coastal West Africa, consistent with the high rates reported in studies not included in this analysis, such as in the Central African Republic^69^, Guinea Bissau^70^, Mozambique^64^ and Madagascar.^71^ Zones of high predicted prevalence also occurred in South America, Caribbean Central America, the Ganges–Brahmaputra Delta, and New Guinea among others, and may reflect the secondary influence of static environmental and household-level risk factors. These findings should inform the design and selection of sites for forthcoming phase 3 *Shigella* vaccine trials^21^, and populations living in these hotspots should be considered high priority for eventual vaccine rollout campaigns.

## Data Availability

The epidemiological data used in this analysis contains identifiable human subject data, which cannot be disseminated under the terms of the IRB and data use agreements with contributing institutions. Investigators from contributing studies may be contacted with reasonable request for data access. Data from the MAL-ED and GEMS studies are available from the ClinEpiDB website.72 GLDAS data is disseminated as part of the mission of NASA's Earth Science Division and archived and distributed by the Goddard Earth Sciences (GES) Data and Information Services Center (DISC).73 Other input data is available from the sources cited in the supplementary appendix. Processed versions in raster/TIFF format are available upon reasonable request to the corresponding author as is statistical source code. Output model predictions of probabilities and standard errors are available for each of nine age groups/symptom stratum combinations at the following GitHub repository https://github.com/joshcolston/Badr_Shigella_predictions.

## Acknowledgements

We would like to thank the many fieldworkers, research teams and participants of the included studies who dedicated their time and information to better the understanding of the transmission and impact of enteric infections in early childhood. GLDAS data is disseminated as part of the mission of NASA’s Earth Science Division and archived and distributed by the Goddard Earth Sciences (GES) Data and Information Services Center (DISC). The findings and conclusions of this report are those of the authors and do not necessarily represent the official position of the authors’ affiliated institutions.

## Author contributions

JMC, MNK and BFZ conceived of the study and obtained ethical approval. JMC and MNK coordinated data sharing. JMC wrote the original draft. MNK and BFZ secured funding for the research and provided review and editing. JMC and HB carried out data processing, analysis, and visualization. NHN carried out literature review. YTC assisted with data visualization. All other authors carried out investigation and provided data sharing support, review, and editing.

## Ethical standards

Ethical approval for this study was given by the University of Virginia Institutional Review Board for Health Sciences Research (IRB-HSR #21544) and the Johns Hopkins University Homewood Institutional Review Board (Study# HIRB0011882). Each contributing study obtained ethical approval from their respective institutions, and written informed consent from participants’ caregivers for collection, storage and analysis of biological specimens as detailed elsewhere. All contributing authors and investigators gave their consent to analyze and publish the data.

## Data Availability Statement

The epidemiological data used in this analysis contains identifiable human subject data, which cannot be disseminated under the terms of the IRB and data use agreements with contributing institutions. Investigators from contributing studies may be contacted with reasonable request for data access. Data from the MAL-ED and GEMS studies are available from the ClinEpiDB website.^72^ GLDAS data is disseminated as part of the mission of NASA’s Earth Science Division and archived and distributed by the Goddard Earth Sciences (GES) Data and Information Services Center (DISC).^73^ Other input data is available from the sources cited in the supplementary appendix. Processed versions in raster/TIFF format are available upon reasonable request to the corresponding author as is statistical source code. Output model predictions of probabilities and standard errors are available for each of nine age groups/symptom stratum combinations at the following GitHub repository https://github.com/joshcolston/Badr_Shigella_predictions.

## References

1 Global Burden of Disease Collaborative Network. Global Burden of Disease Study 2019 (GBD 2019) Results. 2020.

2 Black RE, Brown KH, Becker S. Effects of diarrhea associated with specific enteropathogens on the growth of children in rural Bangladesh. Pediatrics 1984; 73: 799–805.

3 Rogawski ET, Liu J, Platts-Mills JA, et al. Use of quantitative molecular diagnostic methods to investigate the effect of enteropathogen infections on linear growth in children in low-resource settings: longitudinal analysis of results from the MAL-ED cohort study. The Lancet Global Health 2018; 6: e1319–28.

4 Lee G, Paredes Olortegui M, Peñataro Yori P, et al. Effects of Shigella-, Campylobacter- and ETEC-associated diarrhea on childhood growth. Pediatr Infect Dis J 2014; 33: 1004–9.

5 McCormick BJJ, Caulfield LE, Richard SA, et al. Early Life Experiences and Trajectories of Cognitive Development. Pediatrics 2020; 146: e20193660.

6 Bhattacharjee A, Burr AHP, Overacre-Delgoffe AE, et al. Environmental enteric dysfunction induces regulatory T cells that inhibit local CD4+ T cell responses and impair oral vaccine efficacy. Immunity 2021; 54: 1745–1757.e7.

7 DeBoer MD, Chen D, Burt DR, et al. Early childhood diarrhea and cardiometabolic risk factors in adulthood: The Institute of Nutrition of Central America and Panama Nutritional Supplementation Longitudinal Study. Annals of Epidemiology 2013; 23: 314–20.

8 Kim MJ, Moon Y, Kim H, et al. Cross-Protective Shigella Whole-Cell Vaccine With a Truncated O-Polysaccharide Chain. Frontiers in Microbiology 2018; 9. DOI:10.3389/fmicb.2018.02609.

9 Kotloff KL, Winickoff JP, Ivanoff B, et al. Global burden of Shigella infections: implications for vaccine development and implementation of control strategies. Bull World Health Organ 1999; 77: 651–66.

10 Troeger C, Blacker BF, Khalil IA, et al. Estimates of the global, regional, and national morbidity, mortality, and aetiologies of diarrhoea in 195 countries: a systematic analysis for the Global Burden of Disease Study 2016. The Lancet Infectious Diseases 2018; 18: 1211–28.

11 Khalil IA, Troeger C, Blacker BF, et al. Morbidity and mortality due to shigella and enterotoxigenic Escherichia coli diarrhoea: the Global Burden of Disease Study 1990-2016. The Lancet Infectious diseases 2018; 18: 1229–40.

12 Asare EO, Hergott D, Seiler J, et al. Case fatality risk of diarrhoeal pathogens: a systematic review and meta-analysis. International Journal of Epidemiology 2022; : dyac098.

13 Rogawski Mcquade ET, Shaheen F, Kabir F, et al. Epidemiology of Shigella infections and diarrhea in the first two years of life using culture-independent diagnostics in 8 low-resource settings. PLOS Neglected Tropical Diseases 2020; 14: 1–17.

14 Chao DL, Roose A, Roh M, Kotloff KL, Proctor JL. The seasonality of diarrheal pathogens: A retrospective study of seven sites over three years. PLOS Neglected Tropical Diseases 2019; 13: e0007211.

15 Lee HS, Ha Hoang TT, Pham-Duc P, et al. Seasonal and geographical distribution of bacillary dysentery (shigellosis) and associated climate risk factors in Kon Tum Province in Vietnam from 1999 to 2013. Infectious Diseases of Poverty 2017; 6: 113.

16 Colston JM, Faruque ASG, Hossain MJ, et al. Associations between Household-Level Exposures and All-Cause Diarrhea and Pathogen-Specific Enteric Infections in Children Enrolled in Five Sentinel Surveillance Studies. International Journal of Environmental Research and Public Health 2020; 17: 8078.

17 Livio S, Strockbine NA, Panchalingam S, et al. Shigella Isolates From the Global Enteric Multicenter Study Inform Vaccine Development. Clinical Infectious Diseases 2014; 59: 933–41.

18 Hosangadi D, Smith PG, Giersing BK. Considerations for using ETEC and Shigella disease burden estimates to guide vaccine development strategy. Vaccine 2019; 37: 7372.

19 Khalil I, Troeger CE, Blacker BF, Reiner RC. Capturing the true burden of Shigella and ETEC: The way forward. Vaccine 2019; 37: 4784–6.

20 Prudden HJ, Hasso-Agopsowicz M, Black RE, et al. Meeting Report: WHO Workshop on modelling global mortality and aetiology estimates of enteric pathogens in children under five. Cape Town, 28–29th November 2018. In: Vaccine. Elsevier Ltd, 2020: 4792–800.

21 Pavlinac PB, Rogawski McQuade ET, Platts-Mills JA, et al. Pivotal Shigella Vaccine Efficacy Trials-Study Design Considerations from a Shigella Vaccine Trial Design Working Group. Vaccines (Basel) 2022; 10: 489.

22 Kraemer MUG, Hay SI, Pigott DM, Smith DL, Wint GRW, Golding N. Progress and Challenges in Infectious Disease Cartography. Trends in Parasitology. 2016; 32: 19–29.

23 Weiss DJ, Lucas TCD, Nguyen M, et al. Mapping the global prevalence, incidence, and mortality of Plasmodium falciparum, 2000–17: a spatial and temporal modelling study. The Lancet 2019; 394: 322–31.

24 Battle KE, Lucas TCD, Nguyen M, et al. Mapping the global endemicity and clinical burden of Plasmodium vivax, 2000–17: a spatial and temporal modelling study. The Lancet 2019; 394: 332–43.

25 Deshpande A, Miller-Petrie MK, Johnson KB, et al. The global distribution of lymphatic filariasis, 2000–18: a geospatial analysis. The Lancet Global Health 2020; 8: e1186–94.

26 Pigott DM, Bhatt S, Golding N, et al. Global distribution maps of the Leishmaniases. eLife 2014; 2014. DOI:10.7554/eLife.02851.001.

27 Reiner RC, Wiens KE, Deshpande A, et al. Mapping geographical inequalities in childhood diarrhoeal morbidity and mortality in low-income and middle-income countries, 2000–17: analysis for the Global Burden of Disease Study 2017. The Lancet 2020; 395: 1779–801.

28 Bagamian KH, Anderson JD, Muhib F, et al. Heterogeneity in enterotoxigenic Escherichia coli and shigella infections in children under 5 years of age from 11 African countries: a subnational approach quantifying risk, mortality, morbidity, and stunting. The Lancet Global Health 2019; published online Nov. DOI:10.1016/S2214-109X(19)30456-5.

29 Colston JM, Zaitchik BF, Badr HS, et al. Associations Between Eight Earth Observation-Derived Climate Variables and Enteropathogen Infection: An Independent Participant Data Meta-Analysis of Surveillance Studies With Broad Spectrum Nucleic Acid Diagnostics. Geohealth 2022; 6: e2021GH000452.

30 Baker JM, Hasso-Agopsowicz M, Pitzer VE, et al. Association of enteropathogen detection with diarrhoea by age and high versus low child mortality settings: a systematic review and meta-analysis. Lancet Glob Health 2021; 9: e1402–10.

31 Operario DJ, Platts-Mills JA, Nadan S, et al. Etiology of Severe Acute Watery Diarrhea in Children in the Global Rotavirus Surveillance Network Using Quantitative Polymerase Chain Reaction. The Journal of infectious diseases 2017; 216: 220–7.

32 McCormick BJJ, Richard SA, Murray-Kolb LE, et al. Full breastfeeding protection against common enteric bacteria and viruses: results from the MAL-ED cohort study. Am J Clin Nutr 2022; 115: 759–69.

33 Nasrin D, Blackwelder WC, Sommerfelt H, et al. Pathogens associated with linear growth faltering in children with diarrhea and impact of antibiotic treatment: The Global Enteric Multicenter Study. The Journal of Infectious Diseases 2021; published online Sept 16. DOI:10.1093/INFDIS/JIAB434.

34 Rodell M, Houser PR, Jambor U, et al. The Global Land Data Assimilation System. http://dx.doi.org/101175/BAMS-85-3-381 2004.

35 Badr HS. Bindings for Generalized Additive Models (GAM). 2021 https://hsbadr.github.io/additive/ (accessed Nov 22, 2021).

36 Badr HS. Bindings for Bayesian TidyModels. 2021 https://hsbadr.github.io/bayesian/ (accessed Nov 22, 2021).

37 Apley DW, Zhu J. Visualizing the Effects of Predictor Variables in Black Box Supervised Learning Models. 2019; published online Aug 19. DOI:10.48550/arXiv.1612.08468.

38 Štrumbelj E, Kononenko I. Explaining prediction models and individual predictions with feature contributions. Knowl Inf Syst 2014; 41: 647–65.

39 Kraay ANM, Man O, Levy MC, Levy K, Ionides E, Eisenberg JNS. Understanding the impact of rainfall on diarrhea: Testing the concentration-dilution hypothesis using a systematic review and meta-analysis. Environmental Health Perspectives 2020; 128: 126001–1-126001–16.

40 StataCorp. Stata Statistical Software: Release 16. 2019.

41 R Core Team. R: A language and environment for statistical computing. 2020.

42 ESRI. ArcGIS Desktop: Release 10.8. 2019.

43 Stewart LA, Clarke M, Rovers M, et al. Preferred Reporting Items for Systematic Review and Meta-Analyses of individual participant data: the PRISMA-IPD Statement. JAMA 2015; 313: 1657–65.

44 Stevens GA, Alkema L, Black RE, et al. Guidelines for Accurate and Transparent Health Estimates Reporting: the GATHER statement. The Lancet 2016; 388: e19–23.

45 Battle KE, Lucas TCD, Nguyen M, et al. Mapping the global endemicity and clinical burden of Plasmodium vivax, 2000–17: a spatial and temporal modelling study. The Lancet 2019; 394: 332–43.

46 Dwyer-Lindgren L, Cork MA, Sligar A, et al. Mapping HIV prevalence in sub-Saharan Africa between 2000 and 2017. Nature 2019; published online May 15. DOI:10.1038/s41586-019-1200-9.

47 Chammartin F, Scholte RG, Guimarães LH, Tanner M, Utzinger J, Vounatsou P. Soil-transmitted helminth infection in South America: a systematic review and geostatistical meta-analysis. The Lancet infectious diseases 2013; 13: 507–18.

48 Pigott DM, Howes RE, Wiebe A, et al. Prioritising infectious disease mapping. PLoS Neglected Tropical Diseases 2015; 9. DOI:10.1371/journal.pntd.0003756.

49 Hay SI, Battle KE, Pigott DM, et al. Global mapping of infectious disease. Philosophical Transactions of the Royal Society B: Biological Sciences. 2013; 368. DOI:10.1098/rstb.2012.0250.

50 Lanata CF, Black RE. Estimating the true burden of an enteric pathogen: enterotoxigenic Escherichia coli and Shigella spp. The Lancet Infectious Diseases 2018; 18: 1165–6.

51 Hosangadi D, Smith PG, Giersing BK. Considerations for using ETEC and Shigella disease burden estimates to guide vaccine development strategy. Vaccine 2019; 37: 7372.

52 Moyes CL, Temperley WH, Henry AJ, Burgert CR, Hay SI. Providing open access data online to advance malaria research and control. Malaria Journal 2013; 12: 161.

53 Liu J, Ochieng C, Wiersma S, et al. Development of a TaqMan array card for acute-febrile-illness outbreak investigation and surveillance of emerging pathogens, including ebola virus. Journal of Clinical Microbiology 2016; 54: 49–58.

54 Liu J, Gratz J, Amour C, et al. Optimization of Quantitative PCR Methods for Enteropathogen Detection. PLOS ONE 2016; 11: e0158199.

55 Brown J, Cumming O. Perspective Piece Stool-Based Pathogen Detection Offers Advantages as an Outcome Measure for Water, Sanitation, and Hygiene Trials. Am J Trop Med Hyg 2019; 0: 1–2.

56 Operario DJ, Platts-Mills JA, Nadan S, et al. Etiology of Severe Acute Watery Diarrhea in Children in the Global Rotavirus Surveillance Network Using Quantitative Polymerase Chain Reaction. The Journal of infectious diseases 2017; 216: 220–7.

57 Grinberg M, Orevi T, Steinberg S, Kashtan N. Bacterial survival in microscopic surface wetness. eLife 2019; 8. DOI:10.7554/eLife.48508.

58 Chua PLC, Ng CFS, Tobias A, Seposo XT, Hashizume M. Associations between ambient temperature and enteric infections by pathogen: a systematic review and meta-analysis. The Lancet Planetary Health 2022; 6: e202–18.

59 Wang L-P, Zhou S-X, Wang X, et al. Etiological, epidemiological, and clinical features of acute diarrhea in China. Nature Communications 2021; 12: 2464.

60 Zhao Z, Chen Q, Zhao B, et al. Transmission pattern of shigellosis in Wuhan City, China: a modelling study. Epidemiology & Infection 2021; 149. DOI:10.1017/S0950268821002363.

61 Colston J, Paredes Olortegui M, Zaitchik B, et al. Pathogen-Specific Impacts of the 2011–2012 La Niña-Associated Floods on Enteric Infections in the MAL-ED Peru Cohort: A Comparative Interrupted Time Series Analysis. International Journal of Environmental Research and Public Health 2020; 17: 487.

62 Mara D. The elimination of open defecation and its adverse health effects: a moral imperative for governments and development professionals. Journal of Water, Sanitation and Hygiene for Development 2017; 7: 1–12.

63 Saleem M, Burdett T, Heaslip V. Health and social impacts of open defecation on women: a systematic review. BMC Public Health 2019; 19: 158.

64 Knee J, Sumner T, Adriano Z, et al. Effects of an urban sanitation intervention on childhood enteric infection and diarrhea in maputo, mozambique: A controlled before-and-after trial. eLife 2021; 10. DOI:10.7554/ELIFE.62278.

65 Colston JM, Faruque ASG, Hossain MJ, et al. Associations between Household-Level Exposures and All-Cause Diarrhea and Pathogen-Specific Enteric Infections in Children Enrolled in Five Sentinel Surveillance Studies. International Journal of Environmental Research and Public Health 2020; 17: 8078.

66 Vubil D, Acácio S, Quintò L, et al. Clinical features, risk factors, and impact of antibiotic treatment of diarrhea caused by Shigella in children less than 5 years in Manhiça district, rural Mozambique. Infection and Drug Resistance 2018; 11: 2095–106.

67 Kosek M, Yori PP, Pan WK, et al. Epidemiology of highly endemic multiply antibiotic-resistant shigellosis in children in the Peruvian Amazon. Pediatrics 2008; 122: e541–9.

68 Nasrin S, Haque MA, Palit P, et al. Incidence of Asymptomatic Shigella Infection and Association with the Composite Index of Anthropometric Failure among Children Aged 1–24 Months in Low-Resource Settings. Life 2022; 12: 607.

69 Breurec S, Vanel N, Bata P, et al. Etiology and Epidemiology of Diarrhea in Hospitalized Children from Low Income Country: A Matched Case-Control Study in Central African Republic. PLoS Neglected Tropical Diseases 2016; 10. DOI:10.1371/journal.pntd.0004283.

70 Mero S, Timonen S, Lääveri T, et al. Prevalence of diarrhoeal pathogens among children under five years of age with and without diarrhoea in Guinea-Bissau. PLoS Negl Trop Dis 2021; 15: e0009709.

71 Collard J-M, Andrianonimiadana L, Habib A, et al. High prevalence of small intestine bacteria overgrowth and asymptomatic carriage of enteric pathogens in stunted children in Antananarivo, Madagascar. PLoS Negl Trop Dis 2022; 16: e0009849.

72 VEuPathDB. Clinical Epidemiology Resources. ClinEpiDB. 2021. https://clinepidb.org/ce/app (accessed Oct 7, 2021).

73 NASA. Goddard Earth Sciences (GES) Data and Information Services Center (DISC). Earth Data. 2021. https://disc.gsfc.nasa.gov/ (accessed Oct 7, 2021).

