## Supplementary materials for "Spatiotemporal variation in risk of *Shigella* infection in childhood: a global risk mapping and prediction model using individual participant data"

### 1. Training database

### 1.1. Study inclusion criteria

This analysis used an Independent Participant Data Meta Analysis (IPD-MA) framework in which raw, individual participant-level data were pooled from studies that used molecular diagnostics to diagnose *Shigella* in stool samples collected from children aged under 5 years in Low- and Middle-Income Countries (LMICs, as defined by the Organisation for Economic Co-operation and Development [1], excluding those in Europe). The IPD-MA design is considered the gold standard in systematic reviews, offering numerous advantages over aggregate data meta-analyses, and a greater potential for generalizable inferences compared with individual studies [2,3]. In this IPD-MA, studies which tested for *Shigella* using quantitative or real-time polymerase chain reaction (PCR) were identified through non-systematic, exploratory literature review and professional networks and investigators from eligible studies were contacted with requests for access to their data. No specific search strategy was used. If investigators were responsive and agreed to participate, data use agreements were executed between the University of Virginia and the collaborating institution and study-specific datasets were securely transferred and stored, deidentified and combined into a central database with a standardized format and list of variables. Analyses were conducted on the pooled data. **Table S1** summarizes key features of studies that contributed data. No protocol has been registered or published for this study.

| Table S1: Features of studies that contributed to the training database for this analysis | | | | | | | | |
| --- | --- | --- | --- | --- | --- | --- | --- | --- |
| Study | | Sites | Design | Inclusion criteria | Sample collection schedule | Follow-up period | Number of included samples (children) | Reference |
| 1. | Agogo Presbyterian Hospital (APH) | Asante Akim North Municipality, Ghana | Health facility-based unmatched case-control study | Watery/bloody diarrhea cases age <6 years, hospital controls | 1 per child collected within 24 hours of enrollment | 2007-2008 | 646 (606) | [4] |
| 2. | Asian Intussusception Surveillance Network (AISN) | Various locations in Bangladesh, Nepal, Pakistan, and Vietnam | Health facility-based matched case-control study | Intussusception cases age <2 years, matched hospital controls | 1 per child collected within 48 hours of enrollment | 2015-2017 | 393 (393) | [5] |
| 3. | Centre de Recherches Médicales de Lambaréné (CERMEL) | 2 hospitals in Lambaréné, Gabon | Health facility-based surveillance study | Diarrhea patients aged <5 years | 1 per child collected within 48 hours of enrollment | 2017 - 2018 | 179 (179) | [6] |
| 4. | Dar es Salaam Hospitals | 3 major hospitals in Dar es Salaam | Health facility-based unmatched case-control study | Diarrhea cases (any severity) aged <2 years, unmatched hospital controls | 1 per child collected on day of enrollment | 2010-2011 | 1,258 (1,257) | [7] |
| 5. | Delivery of Oral  Cholera Vaccine Effectively (DOVE) project, Cameroon | Douala and the Far North of Cameroon | Health facility-based surveillance study | Acute diarrhea patients aged <18 years | 1 per child collected at enrollment | 2016 - 2018 | 337 (337) | [8] |
| 6. | Diarrhoeal Sentinel Surveillance Programme (DSSP) | Various locations in South Africa | Health facility-based surveillance study | Acute diarrhea patients aged <5 years | 1 per child collected within 48 hours of admission | 2009-2017 | 689 (689) | [9] |
| 7. | Global Enteric Multicenter Study (GEMS) | Mirzapur, Bangladesh; Basse, The Gambia; Kolkota, India; Nyanza, Kenya; Bamako, Mali; Manhiça, Mozambique; Karachi, Pakistan | Health facility-based matched case-control study | Moderate-to-severe diarrhea cases aged <5 years, matched community controls | 1 per child collected at enrollment | 2007-2011 | 11,276 (10,769) | [10] |
| 8. | Haydom Lutheran Hospital (HLH) | Haydom, Tanzania | Health facility-based surveillance study | Gastroenteritis/  diarrhea patients aged <5 years | 1 per child collected within 48 hours of enrollment | 2014-2015 | 233 (233) | [11] |
| 9. | iLiNS-DYAD | Mangochi District, Malawi | Community-based trial of a nutritional product | Children of women enrolled during pregnancy | 3 asymptomatic samples per child collected at 6, 18 and 30 months of age | 2011-2014 | 1,835 (711) | [12] |
| 10. | Livestock-rearing in the Greater Accra Region (GAR) | Ga East and Shai Osudoku Districts of Greater  Accra, Ghana | Community-based cross-sectional study | Children aged 6 – 59 months of caregivers aged ≥18 years | 1 per child collected shortly after enrolment | 2018 | 265 (265) | [13] |
| 11. | Malnutrition and Enteric Disease Study (MAL-ED) | Dhaka, Bangladesh; Fortaleza, Brazil; Vellore, India; Bhaktapur, Nepal; Naushahro Feroze, Pakistan; Loreto, Peru; Venda, South Africa; Haydom, Tanzania | Community-based cohort study | Newborns with normal birth weight | Monthly from 0- 24 months of age and upon caregiver reported diarrhea episode (of any severity) | 2009-2014 | 41,327 (1,715) | [14] |
| 12. | Novel Biomarkers of Environmental Enteropathy (NBEE) | Loreto, Peru | Community-based cohort study | Children aged 3 – 18 months | 15 per child between 1-30 days following enrollment | 2018 | 1,075 (75) | [15] |
| 13. | Programme for Awareness and  Elimination of Diarrhoea (PAED) | Lusaka and Ndola, Zambia | Health facility-based surveillance study | Moderate-to-severe diarrhea patients aged <5 years | 1 per child collected at enrollment | 2012-2013 | 1,379 (1,379) | [16] |
| 14. | Pediatric Hospital of Luanda (PHL) | Luanda, Angola | Health facility-based unmatched case-control study | Diarrhea cases (any severity) aged <5 years, unmatched hospital controls | 1 sample per child collected at enrollment | 2013-2014 | 194 (194) | [17] |
| 15. | RECODISA | Six cities in the semiarid region of Brazil (Cajazeiras, Crato, Ouricuri, Patos, Picos and Sousa) | Community-based matched case-control study | Diarrhea cases (any severity) aged 2-36 months, matched community controls | 1 per child collected at enrollment | 2009-2012 | 1,200 (1,200) | [18] |
| 16. | Rotavac Trial, India | Three sites in India (Delhi, Pune, and Vellore) | Health facility-based placebo-controlled vaccine efficacy trial | Infants aged 6 – 7 weeks | Upon caregiver reported diarrhea episode (any severity) | 2011-2013 | 1,271 (1,090) | [19] |
| 17. | Sanitation Hygiene Infant Nutrition Efficacy (SHINE) Trial | Midlands, Zimbabwe | Community-based WASH intervention trial | HIV-unexposed children of women enrolled during pregnancy | 1, 3, 6, and 12 months of age and upon caregiver reported diarrhea episode (any severity) | 2013-2016 | 2,372 (1,046) | [20] |
| 18. | Thai Hospitals | Various locations in Thailand | Health facility-based matched case-control study | Acute diarrhea cases (any age), matched hospital controls | 1 per child collected at enrollment | 2016-2018 | 473 (473) | [21] |
| 19. | Urban and Rural Guatemala | El Trifinio region, and Guatemala City, Guatemala | Health facility-based trial of a nutritional product | Acute non-bloody diarrhea patients aged 6–35 months | 2 per child collected at enrollment and at study day 31 | 2015-2016 | 585 (299) | [22] |
| 20. | WASH HELPS | Numerous villages in Saravane Province, Lao People’s Democratic Republic (Laos) | Community (school)-based WASH intervention trial | Children <5 in randomly selected households with another child attending primary school | 1 per child collected at enrollment | 2017 | 132 (132) | [23] |

### 1.2. Household- and subject-level covariate data

Most contributing studies collected household- and subject-level data through baseline and follow-up assessments, including numerous variables relevant to enteropathogen infection risk. Covariate ascertainment was not consistent across all studies as some variables were not ascertained in certain studies, while in others information was collected but was incomplete. Furthermore, some anthropometric measurements and feeding status assessments were not contemporaneous with the dates of sample collection. To obtain a complete set of values for all covariates, missing subject-level variable data were interpolated, and household-level variable data imputed. Since some studies lacked data for entire variables, equivalent data was extracted from individual child-level microdata from all standard Demographic and Health Surveys (DHS) [24] and Multiple Indicator Cluster Surveys (MICS) [25] carried out in the same countries dating back to 2005 for the same survey stratum (region and urban/rural status) in which the study sites were located. Survey databases were coded identically to the pooled study database and then appended to it to add more locally relevant information prior to interpolation/imputation.

### 1.2.1. Imputation of missing time-fixed household-level covariate data

Missing data for the household-level covariates were simultaneously imputed using multivariate normal regression (MVN) with an iterative Monte Carlo method with country, survey region, urbanicity, source (survey or study) and linear and quadratic terms for time as additional predictors. These variables were assumed to be static (time-fixed) and, for children for whom they were ascertained at multiple time points during follow-up, the first available value was used, for consistency with those studies that only assessed them at baseline.

### 1.2.2. Interpolation of missing time-varying anthropometric and feeding status data

Missing data relating to the feeding variables were imputed and interpolated using predictions from Cox proportional hazard models that modeled age of first introduction of complementary foods and then of full weaning adjusting for country, survey region, urbanicity, sex, time (with linear and quadratic terms), and source (survey or study). Anthropometric Z-scores were calculated for each child at each available anthropometric assessment based on their length/height, weight and age using the WHO Child Growth Standards STATA igrowup package, with implausible values recoded as missing [26]. Then, linear mixed effects models were fitted to each anthropometric measure in turn (LAZ, WAZ, WHZ) with fixed effects for sex, age (with terms up to fourth order polynomial), time (with linear and quadratic terms), and source (survey or study) and subject- and country-specific random effects. Predictions from these models were used to interpolate missing Z-scores to the child and date of sample collection.

| Table S2: Definitions and sources of covariate variables included in this analysis. | | | |
| --- | --- | --- | --- |
| Variable | Definition | Units/  Categories^1^ | Source |
| Control variables | | | |
| Age | **Training data:** Child’s age at the time of sample collection categorized into three groups | 0-11, 12-23, and 24-59 months | Baseline assessment |
| Symptom status^[1]^ | **Training data:** Stool sample collected during a diarrheal episode (of any severity) or while the child was asymptomatic | Asymptomatic, symptomatic | Study data |
| Study design^1^ | **Training data:** Stool sample collected by a health facility-based or community surveillance study | Community surveillance, health facility | Study data |
| Subject-level covariates | | | |
| Exclusively breastfed | **Training data:** Child exclusively breastfed at time of sample collection (coded “no” if ≥6 months) | Yes, no | Caregiver interview |
|  | **Prediction data:** Prevalence of exclusive breastfeeding among infants <6 months | Proportion | LBD [27], DHS [24], MICS [25] |
| Fully weaned | **Training data:** Child fully weaned (not breastfeeding) at time of sample collection (coded “no” if ≥24 months) | Yes, no | Caregiver interview |
|  | **Prediction data:** Proportion of children <24 months not currently breastfeeding. | Proportion | DHS [24], MICS [25] |
| Stunting | **Training data:** Child moderately-to-severely stunted (Length-for-age Z-score<=-2.0) at time of sample collection | Yes, no | Anthropometric assessment |
|  | **Prediction data:** Prevalence of moderate-to-severe stunting in children <5 years | Proportion | LBD [28], DHS [24], MICS [25] |
| Underweight | **Training data:** Child moderately-to-severely underweight (Weight-for-age Z-score<=-2.0) at time of sample collection | Yes, no | Anthropometric assessment |
|  | **Prediction data:** Prevalence of moderate-to-severe underweight in children <5 years | Proportion | LBD [28], DHS [24], MICS [25] |
| Wasting | **Training data:** Child moderately-to-severely wasted (Weight-for-length Z-score<=-2.0) at time of sample collection | Yes, no | Anthropometric assessment |
|  | **Prediction data:** Prevalence of moderate-to-severe wasting in children <5 years | Proportion | LBD [28], DHS [24], MICS [25] |
| Household-level covariates | | | |
| Floor material | **Training data:** Child resides in a household with improved (rudimentary or finished) vs. unimproved (bare earth, sand) floors [29] | Improved, unimproved | Baseline assessment |
|  | **Prediction data:** Proportion of households with improved flooring | Proportion | DHS [24], MICS [25] |
| Household crowding | **Training data:** Child resides in a household with >=3 residents per sleeping room [30] | Yes, no | Baseline assessment |
|  | **Prediction data:** Proportion of households with >=3 residents per sleeping room | Proportion | DHS [24], MICS [25] |
| Caregiver education | **Training data:** Child’s primary caregiver completed primary (6 years of) education | Yes, no | Baseline assessment |
|  | **Prediction data:** Proportion of female population 15-49 that have completed primary (6 years of) education | Proportion | LBD [31], DHS [24], MICS [25] |
| Open defecation | **Training data:** Child resides in a household that practices open defecation (has no sanitation facility) [32] | Yes, no | Baseline assessment |
|  | **Prediction**: Proportion of households practicing open defecation (no sanitation facility) | Proportion | LBD [33], DHS [24], MICS [25] |
| Sanitation facility | **Training data:** Child resides in a household with an improved vs. unimproved sanitation facility [32] | Improved, unimproved | Baseline assessment |
|  | **Prediction**: Proportion of households with an improved sanitation facility | Proportion | LBD [33], DHS [24], MICS [25] |
| Water source | **Training data:** Child resides in a household with an improved vs. unimproved drinking water source [32] | Improved, unimproved | Baseline assessment |
|  | **Prediction**: Proportion of households with an improved sanitation facility | Proportion | LBD [33], DHS [24], MICS [25] |
| Environmental spatial covariates | | | |
| Accessibility to cities | Travel time to nearest settlement of >50,000 inhabitants | Minutes | MAP [34] global raster |
| Cropland areas | Proportion of land given over to cropland, 2000 | Proportion | CIESIN [35] global raster |
| Distance to major river | Distance to major perennial river (derived from rivers and lakes centerlines database) | Decimal degrees | Natural Earth [36] global raster |
| Elevation | Elevation above sea level | Meters | NOAA [37] global raster |
| Enhanced Vegetation Index | Vegetation greenness corrected for atmospheric conditions and canopy background noise | Ratio | USGS [38] global raster |
| Growing season length | Reference length of annual agricultural growing period (baseline period 1961-1990) | Days | FAO, IIASA [39] global raster |
| Human Footprint Index | Human Influence Index (HII) normalized by biome and realm. | Percentage | CIESIN [40] global raster |
| Irrigated areas | Percentage of land equipped for irrigation around the year, 2000 | Percentage | FAO [41], global raster |
| Population density | Human population density per 1km^2^ | Inhabitants per km^2^ | WorldPop [42] global raster |
| Urbanicity | Urbanicity status at georeferenced location (reclassified from Global Human Settlement database) | Urban, peri-urban, rural | GHS [43] global raster |
| Time-varying hydrometeorological variables | | | |
| Precipitation deviations | Deviations from the mean daily precipitation in the 7-day period from *t*_-9_ to *t*_-3_ days | Millimeters | GLDAS [44] |
| Relative humidity | Average daily relative humidity in the 7-day period from *t*_-9_ to *t*_-3_ days | Percentage | GLDAS [44] |
| Soil moisture | Average daily soil moisture in the 7-day period from *t*_-9_ to *t*_-3_ days | Percentage | GLDAS [44] |
| Solar radiation | Average daily solar radiation in the 7-day period from *t*_-9_ to *t*_-3_ days | Watts per square meter | GLDAS [44] |
| Specific humidity | Average daily specific humidity in the 7-day period from *t*_-9_ to *t*_-3_ days | Grams/kilogram | GLDAS [44] |
| Surface pressure deviations | Deviations from the mean daily surface pressure in the 7-day period from *t*_-9_ to *t*_-3_ days | Millibars | GLDAS [44] |
| Surface runoff | Average daily surface runoff in the 7-day period from *t*_-9_ to *t*_-3_ days | Millimeters | GLDAS [44] |
| Temperature | Average daily temperature in the 7-day period from *t*_-9_ to *t*_-3_ days | Degrees Celsius | GLDAS [44] |
| Wind speed | Average daily wind speed in the 7-day period from *t*_-9_ to *t*_-3_ days | Meters per second | GLDAS [44] |
| ^1^ For categorical variables, the reference category is underlined.  LBD – IHME’s Local Burden of Disease project; DHS – Demographic and Health Surveys; MICS – Multiple Indicator Cluster Surveys; MAP – Malaria Atlas Project; CIESIN - Center for International Earth Science Information Network; NOAA – National Oceanic and Atmospheric Administration; FAO – Food and Agriculture Organization; IIASA – International Institute for Applied Systems Analysis; GHS – Global Human Settlements; GLDAS – Global Land Data Assimilation System. | | | |

| Table S3: Percent of children with available household- and subject-level variables prior to interpolation and imputation by study | | | | | | | | | |
| --- | --- | --- | --- | --- | --- | --- | --- | --- | --- |
| Study | Household-level variables | | | | | Subject-level variables | | | |
|  | Crowding | Flooring material | Caregiver education | Sanitation facility | Water source | Exclus-ively breastfed | Fully weaned | Height/  length | Weight |
| AISN | 0.0 | 0.0 | 100.0 | 0.0 | 0.0 | 42.7 | 22.6 | 0.0 | 0.0 |
| APH | 0.0 | 18.1 | 17.9 | 18.1 | 18.1 | 17.4 | 62.2 | 0.0 | 0.0 |
| CERMEL | 0.0 | 0.0 | 0.0 | 47.3 | 48.8 | 53.1 | 89.3 | 0.0 | 0.0 |
| DOVE | 0.0 | 0.0 | 0.0 | 0.0 | 21.8 | 0.0 | 0.0 | 0.0 | 0.0 |
| DSSP | 0.0 | 0.0 | 78.8 | 95.7 | 95.5 | 100.0 | 100.0 | 0.0 | 66.5 |
| Dar es Salaam Hospitals | 0.0 | 0.0 | 100.0 | 0.0 | 0.0 | 24.2 | 24.2 | 86.0 | 98.8 |
| GAR | 0.0 | 100.0 | 93.2 | 100.0 | 100.0 | 100.0 | 100.0 | 99.6 | 100.0 |
| GEMS | 100.0 | 100.0 | 100.0 | 100.0 | 100.0 | 81.2 | 81.2 | 100.0 | 100.0 |
| HLH | 0.0 | 0.0 | 0.0 | 0.0 | 0.0 | 0.0 | 0.0 | 0.0 | 0.0 |
| iLiNS-DYAD | 0.0 | 0.0 | 95.3 | 95.6 | 95.4 | 52.9 | 52.8 | 86.4 | 88.7 |
| MAL-ED | 91.4 | 91.4 | 91.4 | 91.4 | 91.4 | 99.4 | 99.4 | 100.0 | 100.0 |
| NBEE | 98.7 | 100.0 | 94.7 | 100.0 | 100.0 | 100.0 | 100.0 | 100.0 | 100.0 |
| PAED | 0.0 | 95.7 | 95.8 | 85.0 | 91.0 | 57.5 | 62.7 | 35.5 | 8.0 |
| PHL | 0.0 | 0.0 | 0.0 | 90.7 | 86.1 | 84.5 | 84.5 | 84.0 | 97.4 |
| RECODISA | 0.0 | 99.3 | 99.7 | 100.0 | 99.8 | 100.0 | 100.0 | 99.8 | 99.8 |
| Rotavac Trial | 0.0 | 0.0 | 0.0 | 0.0 | 0.0 | 0.0 | 0.0 | 0.0 | 0.0 |
| SHINE | 0.0 | 94.7 | 97.7 | 94.2 | 94.5 | 82.7 | 82.5 | 96.7 | 99.9 |
| Thai Hospitals | 0.0 | 0.0 | 0.0 | 0.0 | 0.0 | 0.0 | 0.0 | 0.0 | 0.0 |
| Urban & Rural Guatemala | 0.0 | 0.0 | 0.0 | 100.0 | 100.0 | 0.0 | 0.0 | 100.0 | 100.0 |
| WASH Helps | 0.0 | 100.0 | 100.0 | 100.0 | 100.0 | 0.0 | 0.0 | 0.0 | 0.0 |

### 1.3. Georeferencing and spatial covariate extraction:

For environmental spatial covariate extraction, stool samples were georeferenced to the approximate location of the child’s residence. For some study sites, the exact coordinates of the child’s household locations were available, otherwise, children were georeferenced to the approximate centroid of their neighborhood, village, or district or, where such information was unavailable, the location of the health facility at which they were enrolled. For samples georeferenced to health facilities, covariates were averaged over a theoretical catchment area represented by a 20km buffer around the facility location using the ArcMap Zonal Statistics tool, otherwise they were extracted to household or community locations using the Extract Values to Points tool [45].

### 1.4. Hydrometeorological variable extraction, standardization, and lagging:

The effect of weather on enteropathogen outcomes is lagged [46,47] and the incubation period for shigellosis can be as short as 12 hours, with an average interval of three days from exposure [48] Therefore, daily hydrometeorological variables were averaged over a 7-day lagged period of exposure from 3 to 9 days prior to the date of sample collection (*t*_-9_ to *t*_-3_, where *t_0_* is the date of sample collection). Two of the hydrometeorological exposure variables – precipitation and surface pressure - were standardized to their local distributions by recalculating each one as the deviation from its site-specific mean value over the period 2005 – 2019. For precipitation, this was to account for the distribution being highly right-skewed, while for surface pressure, which was narrowly distributed within each site, it reduced between-site relative to within-site variability [46].

### 2. Prediction databases

### 2.1. Creation of covariate rasters:

To predict *Shigella* infection probability from the model across all LMICs, it was necessary either to obtain raster datasets of the values for each covariate predictor that are coextensive with that domain, or to fix the prediction at a certain value of that variables. For the hydrometeorological and static spatial covariates, rasters are available covering the entire domain of interest. For several of the household- and subject-level variables, raster predictions are available from IHME’s Local Burden of Disease (LBD) project website [33], though most do not cover the entire domain. For variables for which such rasters were available, predictions for the most recent year (2017) were used. For other such variables, and to fill gaps in coverage of the LBD rasters, subnational unit-level values were calculated using microdata from each LMIC’s most recent nationally representative household survey (Standard DHS, MICS or, in small number of cases, country-specific surveys [49,50]), merged with unit polygon shapefiles and converted to raster format. For countries for which no eligible surveys were available in the public domain (e.g., North Korea, Eritrea, Iran), averages across all other countries in that country’s World Bank region [51] were substituted. All covariate rasters were clipped and resampled to match the extent and resolution of the GLDAS grids. **Figures S1**, **S2**, and **S3** show the geographical variation in, respectively, averages of the hydrometeorological variables, the time-static environmental spatial covariates, and the subject- and household-level covariates.

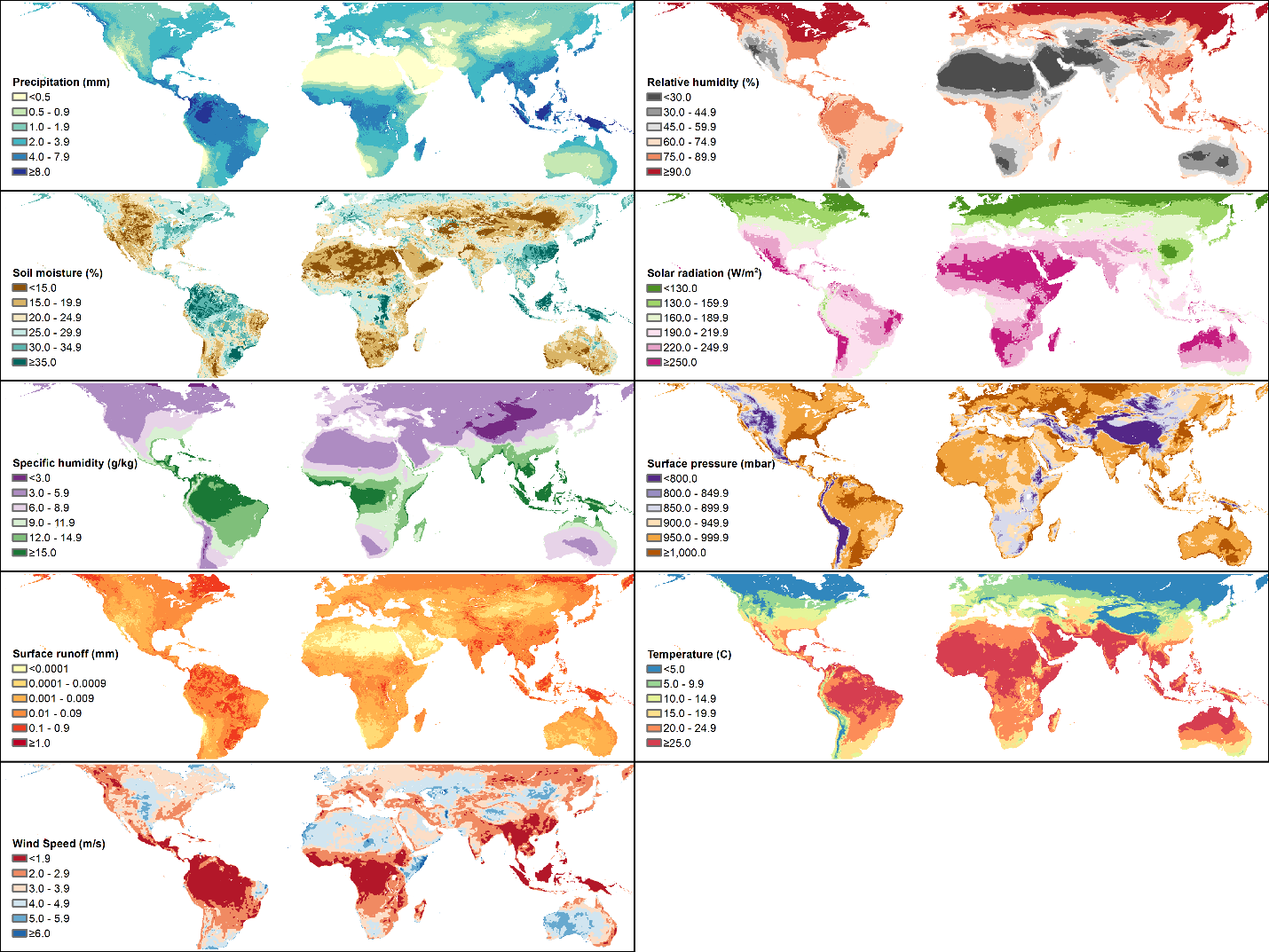

**Figure S1:** Geographical variation in the 9 hydrometeorological variables used in this analysis – average of all values from 2005 – 2019.

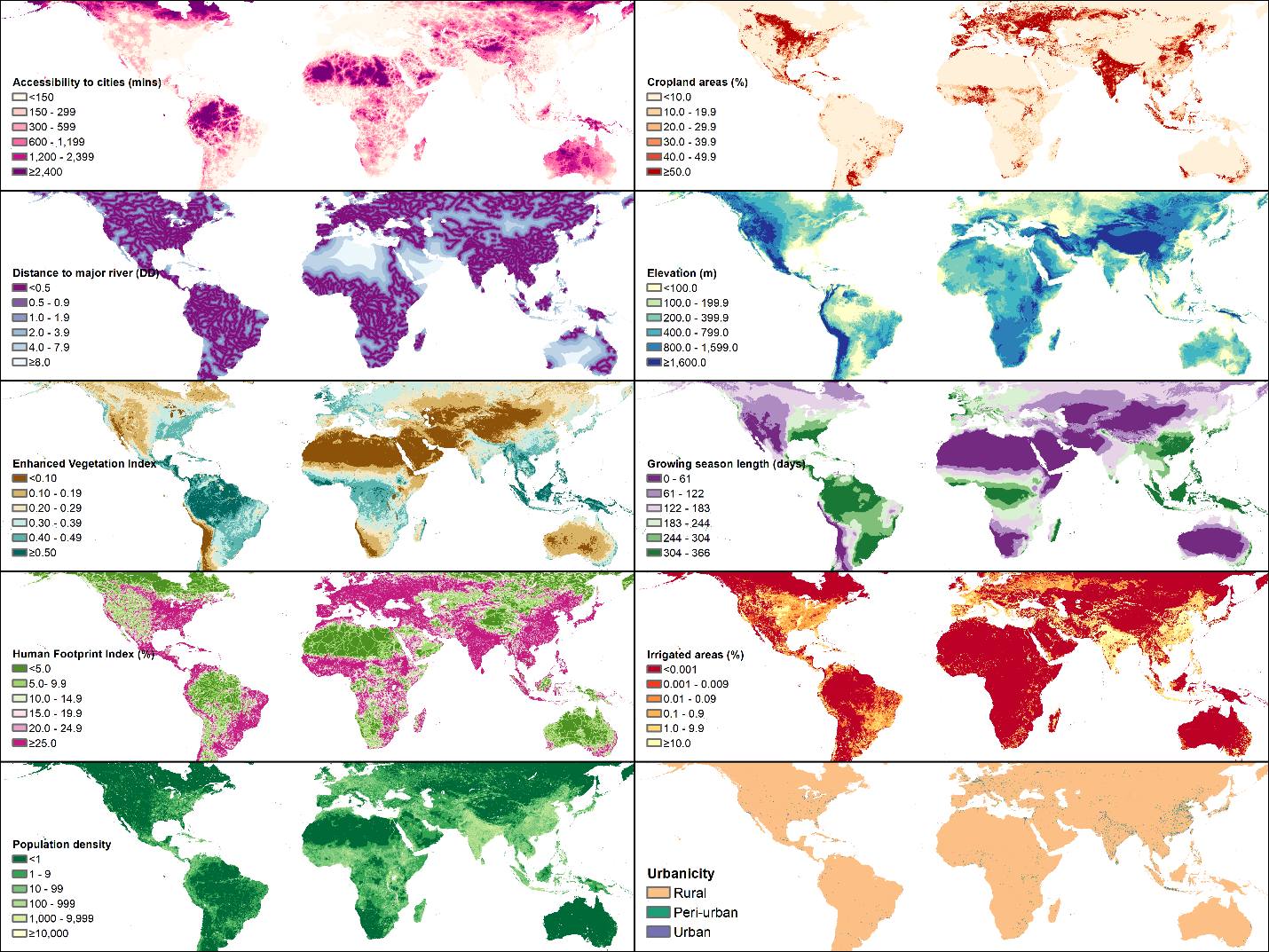

**Figure S2:** Geographical variation in the 8 static environmental covariates used in the analysis.

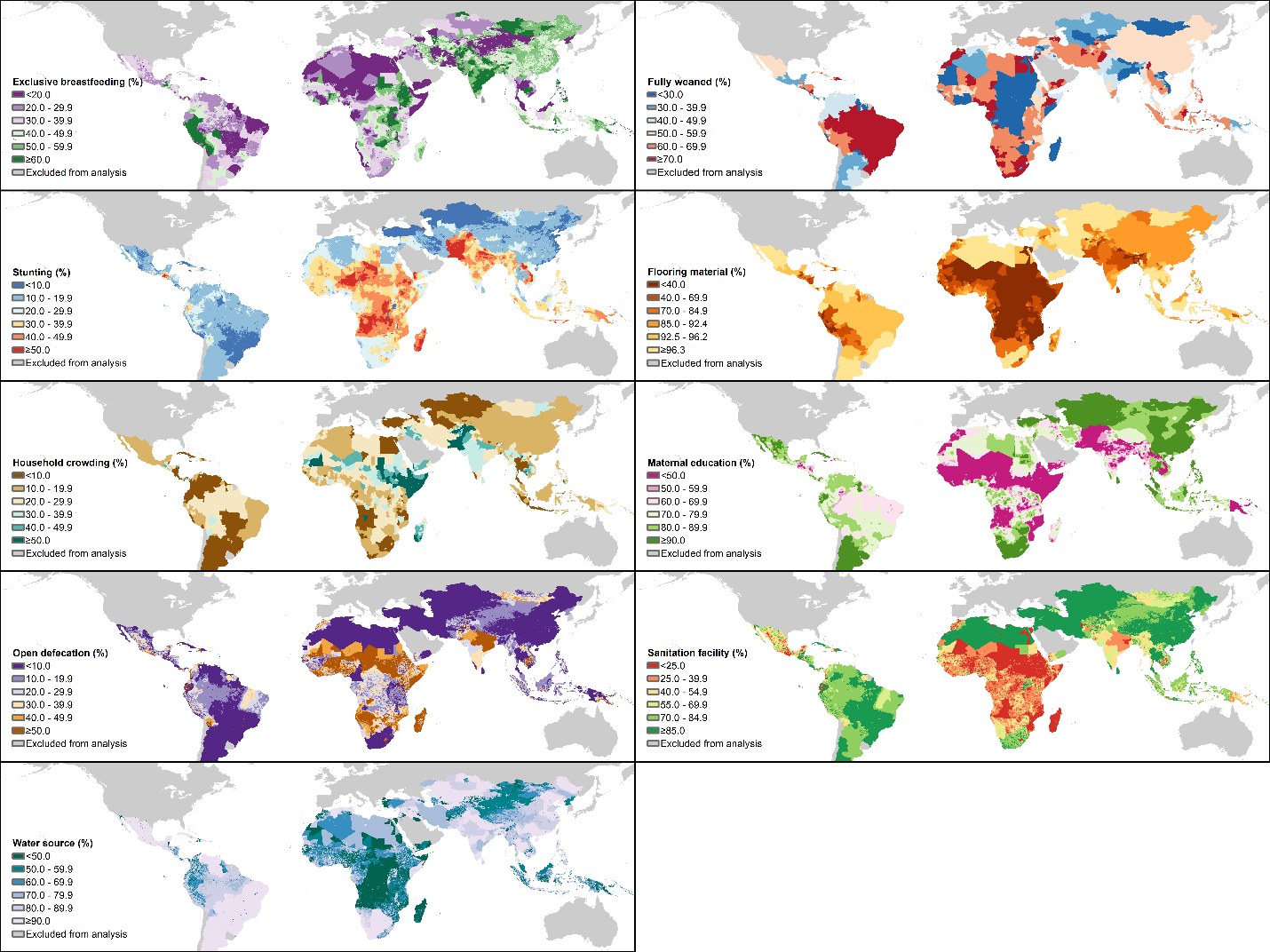

**Figure S3**: Geographical variation in 3 subject-level and 6 household-level covariates used in the analysis.

### 3. Model equation:

**Final model equation S1:**

$$\begin{matrix} \log\left[ \frac{P\left( Positive Shigella \right)}{1-P\left( Positive Shigella \right)} \right] & =\alpha+\beta_{1}\left( {\mathrm{Child}^{'}s age}_{\left[ 12, 24 \right)} \right)+\beta_{2}\left( {\mathrm{Child}^{'}s age}_{\left[ 24, 60 \right)} \right)+\beta_{3}\left( {Symptom status}_{Symptomatic\_CS} \right) +\beta_{4}\left( {Symptom status}_{Symptomatic\_HF} \right)+ \\ & \beta_{5}\left( Exclusively breastfed \right)+\beta_{6}\left( Fully weaned \right)+\beta_{7}\left( \mathrm{Stunting} \right)+\beta_{8}\left( \mathrm{Underweight} \right) +\beta_{9}\left( \mathrm{Wasting} \right)+ \\ & \beta_{10}\left( Household crowding \right)+\beta_{11}\left( Caregiver education \right)+\beta_{12}\left( Floor material \right) +\beta_{13}\left( Open defecation \right)+ \\ & \beta_{14}\left( Sanitation facility \right)+\beta_{15}\left( Water source \right)+\beta_{16}\left( Accessibility to cities \right) +\beta_{17}\left( Cropland areas \right)+ \\ & \beta_{18}\left( Population density \right)+\beta_{19}\left( \mathrm{Elevation} \right)+\beta_{20}\left( Enhanced vegetation index \right) +\beta_{21}\left( Human footprint index \right)+ \\ & \beta_{22}\left( Growing season length \right)+\beta_{23}\left( Irrigated areas \right)+\beta_{24}\left( Distance to major river \right) +\beta_{25}\left( \mathrm{Urbanicity} \right) + \\ & f_{1}\left( Surface pressure \right)+f_{2}\left( Relative humidity \right)+f_{3}\left( Surface runoff \right)+f_{4}\left( Specific humidity \right)+ \\ & f_{5}\left( Solar radiation \right) +f_{6}\left( Wind speed \right)+f_{7}\left( Air temperature, Symptom status \right)+f_{8}\left( Soil moisture, Precipitation \right) \\ & \\ & \end{matrix}$$

Where $\beta_{i}$ represent the parametric coefficients of each variable, while $f_{i}$ are spline smooth functions of the hydrometeorological variables or their interactions. CS = community surveillance study design; HF = health facility-based study design.

### 4. Supplementary results:

Output model predictions of probabilities and standard errors are available for each of nine age groups/symptom stratum combinations at the following GitHub repository <https://github.com/joshcolston/Badr_Shigella_predictions>.

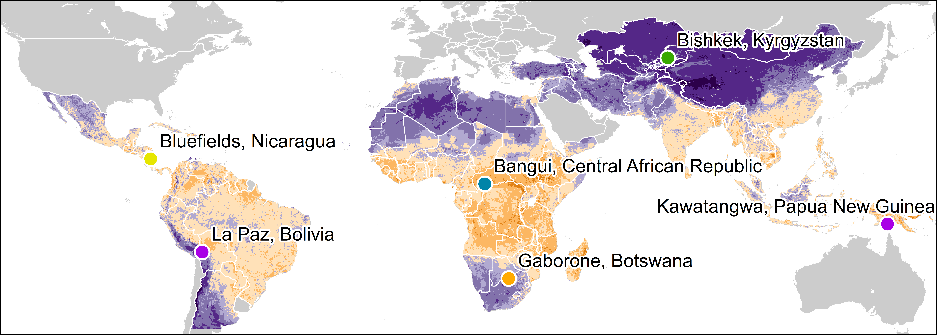

**a).**

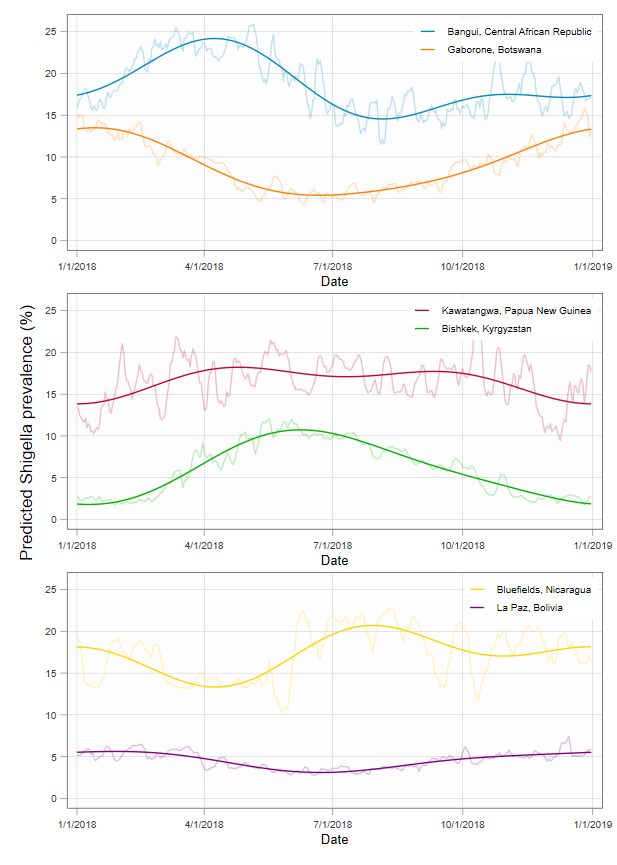

**b).**

**c).**

**d).**

**Figure S4:** Time series plots of the daily (transparent) and smoothed (opaque) estimates of *Shigella* prevalence in asymptomatic children aged 12-23 months over 2018 at six illustrative high and lower prevalence locations (a.) in in Africa (b.), Asia (c.) and the Americas (d.). Smoothing was done using regression with annual and biannual harmonic terms [52]

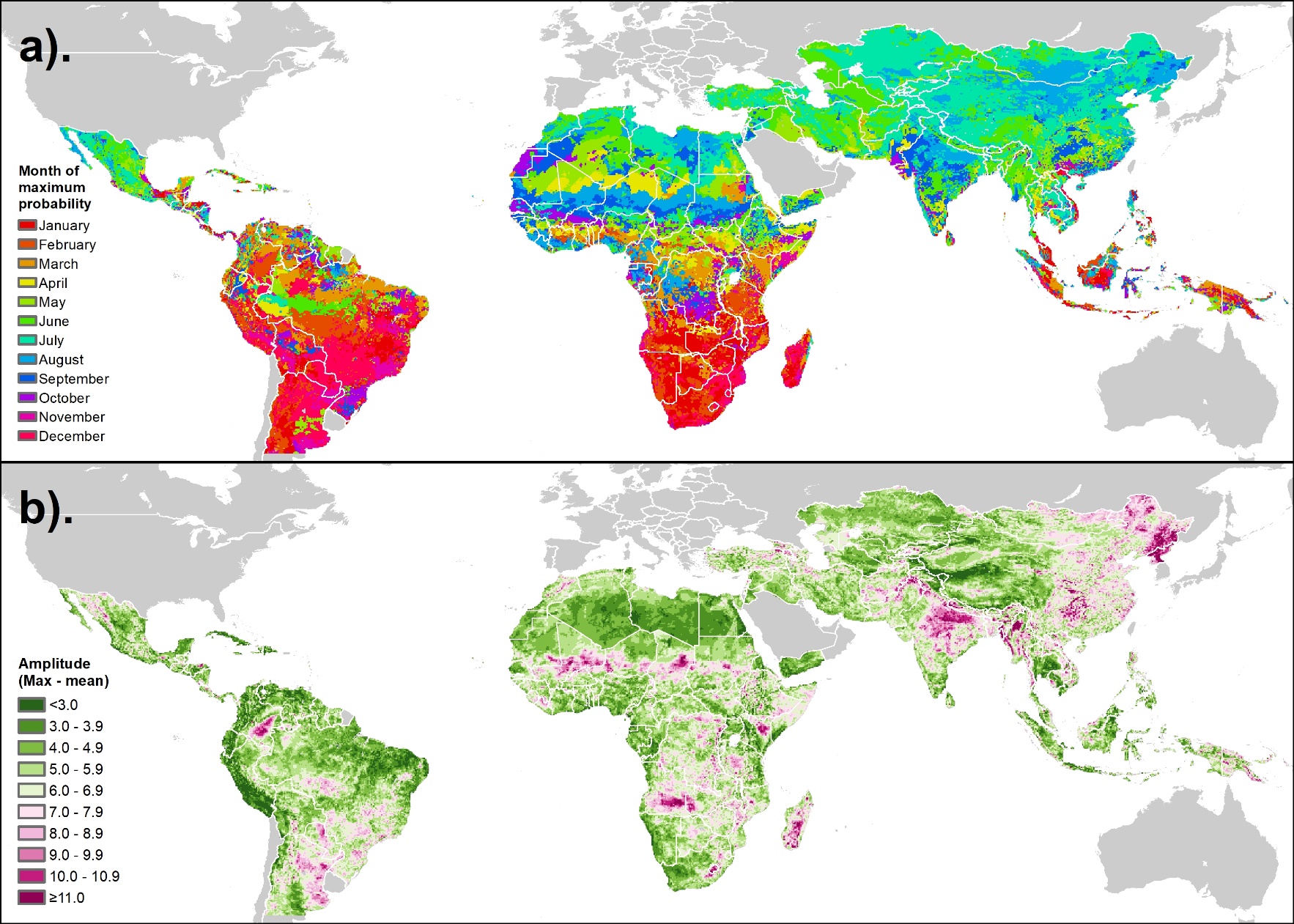

**Figure S5:** Seasonality metrics for *Shigella* positivity in asymptomatic children aged 12-23 months predicted by the model: a). Timing of annual peak (month in which maximum daily predicted positivity in 2018 occurs); b) Amplitude of annual peak (percentage point difference between the maximum and mean daily values in 2018)

### 6. Guideline compliance

| Table S4: Checklist for Guidelines for Accurate and Transparent Health Estimates Reporting (GATHER) [53] | |
| --- | --- |
| Objectives and funding | |
| 1. Define the indicator(s), populations (including age, sex, and geographic entities), and time period(s) for which estimates were made. | Methods - objectives and scope section |
| 2. List the funding sources for the work | Financial support section |
| Data inputs | |
| 3. Describe how the data were identified and how the data were accessed. | Methods section and supplementary tables S1 and S2 |
| 4. Specify the inclusion and exclusion criteria. Identify all ad-hoc exclusions. | Methods section and supplementary appendix section 1.1 |
| 5. Provide information on all included data sources and their main characteristics. For each data source used, report reference information or contact name/institution, population represented, data collection method, year(s) of data collection, sex and age range, diagnostic criteria or measurement method, and sample size, as relevant. | Methods section and supplementary tables S1 and S2 |
| 6. Identify and describe any categories of input data that have potentially important biases (e.g., based on characteristics listed in item 5). | Methods section and supplementary appendix sections 1 and 2 |
| 7. Describe and give sources for any other data inputs. | Not applicable |
| 8. Provide all data inputs in a file format from which data can be efficiently extracted (e.g., a spreadsheet rather than a PDF), including all relevant meta-data listed in item 5. For any data inputs that cannot be shared because of ethical or legal reasons, such as third-party ownership, provide a contact name or the name of the institution that retains the right to the data. | See data availability statement. |
| Data analysis | |
| 9. Provide a conceptual overview of the data analysis method. A diagram may be helpful. | Methods – statistical analysis section |
| 10. Provide a detailed description of all steps of the analysis, including mathematical formulae. This description should cover, as relevant, data cleaning, data pre-processing, data adjustments and weighting of data sources, and mathematical or statistical model(s). | Methods – statistical analysis section; Supplementary equation S1 |
| 11. Describe how candidate models were evaluated and how the final model(s) were selected. | Methods – statistical analysis section |
| 12. Provide the results of an evaluation of model performance, if done, as well as the results of any relevant sensitivity analysis. | Figure 2 |
| 13. Describe methods of calculating uncertainty of the estimates. State which sources of uncertainty were, and were not, accounted for in the uncertainty analysis. | Methods – statistical analysis section |
| 14. State how analytical or statistical source code used to generate estimates can be accessed. | See data availability statement. |
| Results and discussion | |
| 15. Provide published estimates in a file format from which data can be efficiently extracted. | Supplementary TIFF files of estimates probabilities |
| 16. Report a quantitative measure of the uncertainty of the estimates (e.g., uncertainty intervals). | Supplementary TIFF files of standard errors of estimates |
| 17. Interpret results in light of existing evidence. If updating a previous set of estimates, describe the reasons for changes in estimates. | Discussion section |
| 18. Discuss limitations of the estimates. Include a discussion of any modelling assumptions or data limitations that affect interpretation of the estimates. | Discussion section |

| Table S5: Checklist for reporting Systematic Reviews and Meta-Analyses of Individual Participant Data [54] | | |
| --- | --- | --- |
| Title | | |
| 1. | Identify the report as a systematic review and meta-analysis of individual participant data. | Title |
| Abstract | | |
| 2. | Provide a structured summary including as applicable: |  |
|  | **Background:** state research question and main objectives, with information on participants, interventions, comparators, and outcomes. | Abstract |
|  | **Methods:** report eligibility criteria; data sources including dates of last bibliographic search or elicitation, noting that IPD were sought; methods of assessing risk of bias. | Not applicable |
|  | **Results:** provide number and type of studies and participants identified and number (%) obtained; summary effect estimates for main outcomes (benefits and harms) with confidence intervals and measures of statistical heterogeneity. Describe the direction and size of summary effects in terms meaningful to those who would put findings into practice. | Not applicable |
|  | **Discussion:** state main strengths and limitations of the evidence, general interpretation of the results and any important implications. | Abstract |
|  | **Other:** report primary funding source, registration number and registry name for the systematic review and IPD meta-analysis. | Abstract |
| Introduction | | |
| 3. | Describe the rationale for the review in the context of what is already known. | Introduction |
| 4. | Provide an explicit statement of the questions being addressed with reference, as applicable, to participants, interventions, comparisons, outcomes, and study design (PICOS). Include any hypotheses that relate to particular types of participant-level subgroups. | Introduction |
| Method | | |
| 5. | Indicate if a protocol exists and where it can be accessed. If available, provide registration information including registration number and registry name. Provide publication details, if applicable. | Supplementary appendix section 1.1 |
| 6. | Specify inclusion and exclusion criteria including those relating to participants, interventions, comparisons, outcomes, study design and characteristics (e.g., years when conducted, required minimum follow-up). Note whether these were applied at the study or individual level i.e., whether eligible participants were included (and ineligible participants excluded) from a study that included a wider population than specified by the review inclusion criteria. The rationale for criteria should be stated. | Methods and supplementary appendix section 1.1 |
| 7. | Describe all methods of identifying published and unpublished studies including, as applicable: which bibliographic databases were searched with dates of coverage; details of any hand searching including of conference proceedings; use of study registers and agency or company databases; contact with the original research team and experts in the field; open adverts and surveys. Give the date of last search or elicitation. | Supplementary appendix section 1.1 |
| 8. | Present the full electronic search strategy for at least one database, including any limits used, such that it could be repeated. | Not applicable |
| 9. | State the process for determining which studies were eligible for inclusion. | Supplementary appendix section 1.1 |
| 10. | Describe how IPD were requested, collected, and managed, including any processes for querying and confirming data with investigators. If IPD were not sought from any eligible study, the reason for this should be stated (for each such study). | Supplementary appendix section 1.1 |
|  | If applicable, describe how any studies for which IPD were not available were dealt with. This should include whether, how and what aggregate data were sought or extracted from study reports and publications (such as extracting data independently in duplicate) and any processes for obtaining and confirming these data with investigators. | Not applicable. |
| 11. | Describe how the information and variables to be collected were chosen. List and define all study level and participant level data that were sought, including baseline and follow-up information. If applicable, describe methods of standardizing or translating variables within the IPD datasets to ensure common scales or measurements across studies. | Supplementary appendix section 1, supplementary tables S1 & S2 |
|  | Describe what aspects of IPD were subject to data checking (such as sequence generation, data consistency and completeness, baseline imbalance) and how this was done. | Not applicable. |
| 12. | Describe methods used to assess risk of bias in the individual studies and whether this was applied separately for each outcome. If applicable, describe how findings of IPD checking were used to inform the assessment. Report if and how risk of bias assessment was used in any data synthesis. | Not applicable. |
| 13. | State all treatment comparisons of interests. State all outcomes addressed and define them in detail. State whether they were pre-specified for the review and, if applicable, whether they were primary/main or secondary/additional outcomes. Give the principal measures of effect (such as risk ratio, hazard ratio, difference in means) used for each outcome. | Not applicable. |
| 14. | Describe the meta-analysis methods used to synthesize IPD. Specify any statistical methods and models used. | Supplementary appendix section 2 |
|  | If applicable, describe any methods used to explore variation in effects by study or participant level characteristics. | Not applicable. |
| 15. | Specify any assessment of risk of bias relating to the accumulated body of evidence, including any pertaining to not obtaining IPD for studies, outcomes or other variables. | Methods, Covariates section |
| 16. | Describe methods of any additional analyses, including sensitivity analyses. State which of these were pre-specified. | Methods, Statistical Analysis section. |
| Results | | |
| 17. | Give numbers of studies screened, assessed for eligibility, and included in the systematic review with reasons for exclusions at each stage. Indicate the number of studies and participants for which IPD were sought and for which IPD were obtained. For those studies where IPD were not available, give the numbers of studies and participants for which aggregate data were  available. Report reasons for non-availability of IPD. Include a flow diagram. | Not appropriate. |
| 18. | For each study, present information on key study and participant characteristics (such as description of interventions, numbers of participants, demographic data, unavailability of outcomes, funding source, and if applicable duration of follow-up). Provide (main) citations for each study. Where applicable, also report similar study characteristics for any studies not providing IPD | Supplementary table S1 |
|  | Report any important issues identified in checking IPD or state that there were none. | Not applicable |
| 19. | Present data on risk of bias assessments. If applicable, describe whether data checking led to the up-weighting or down-weighting of these assessments. Consider how any potential bias impacts on the robustness of meta-analysis conclusions. | Methods, Data sources and outcome variable section; Discussion |
| 20. | For each comparison and for each main outcome (benefit or harm), for each individual study report the number of eligible participants for which data were obtained and show simple summary data for each intervention group (including, where applicable, the number of events), effect estimates and confidence intervals. These may be tabulated or included on a forest plot. | Figure 1 |
| 21. | Present summary effects for each meta-analysis undertaken, including confidence intervals and measures of statistical heterogeneity. State whether the analysis was pre-specified, and report the numbers of studies and participants and, where applicable, the number of events on which it is based. | Results, figure 3, supplementary table S1 |
|  | When exploring variation in effects due to patient or study characteristics, present summary interaction estimates for each characteristic examined, including confidence intervals and measures of statistical heterogeneity. State whether the analysis was pre-specified. State whether any interaction is consistent across trials. | Not applicable |
|  | Provide a description of the direction and size of effect in terms meaningful to those who would put findings into practice. | Results, figure 3 |
| 22. | Present results of any assessment of risk of bias relating to the accumulated body of evidence, including any pertaining to the availability and representativeness of available studies, outcomes or other variables. | Effect estimate of study design, figure 3 |
| 23. | Give results of any additional analyses (e.g., sensitivity analyses). If applicable, this should also include any analyses that incorporate aggregate data for studies that do not have IPD. If applicable, summarize the main meta-analysis results following the inclusion or exclusion of studies for which IPD were not available. | Not applicable |
| Discussion | | |
| 24. | Summarize the main findings, including the strength of evidence for each main outcome. | Discussion |
| 25. | Discuss any important strengths and limitations of the evidence including the benefits of access to IPD and any limitations arising from IPD that were not available. | Discussion |
| 26. | Provide a general interpretation of the findings in the context of other evidence. | Discussion |
|  | Consider relevance to key groups (such as policy makers, service providers and service users). Consider implications for future research. | Discussion |
| Funding | | |
| 27. | Describe sources of funding and other support (such as supply of IPD), and the role in the systematic review of those providing such support. | Abstract and financial support section |

### 7. References:

1.	Organisation for Economic Co-operation and Development. DAC List of ODA Recipients. In: OECD [Internet]. 2020 [cited 10 Dec 2021]. Available: https://www.oecd.org/dac/financing-sustainable-development/development-finance-standards/daclist.htm

2.	Chen B, Benedetti A. Quantifying heterogeneity in individual participant data meta-analysis with binary outcomes. Systematic Reviews. 2017;6. doi:10.1186/s13643-017-0630-4

3.	Dewidar O, Riddle A, Ghogomu E, Hossain A, Arora P, Bhutta ZA, et al. PRIME-IPD SERIES Part 1. The PRIME-IPD tool promoted verification and standardization of study datasets retrieved for IPD meta-analysis. Journal of Clinical Epidemiology. 2021. doi:10.1016/j.jclinepi.2021.05.007

4.	Eibach D, Krumkamp R, Hahn A, Sarpong N, Adu-Sarkodie Y, Leva A, et al. Application of a multiplex PCR assay for the detection of gastrointestinal pathogens in a rural African setting. BMC Infectious Diseases. 2016;16: 150. doi:10.1186/s12879-016-1481-7

5.	Burnett E, Kabir F, Van Trang N, Rayamajhi A, Satter SM, Liu J, et al. Infectious Etiologies of Intussusception among Children <2 Years Old in 4 Asian Countries. Journal of Infectious Diseases. 2020;221: 1499–1505. doi:10.1093/infdis/jiz621

6.	Manouana GP, Byrne N, Mbong Ngwese M, Nguema Moure A, Hofmann P, Bingoulou Matsougou G, et al. Prevalence of Pathogens in Young Children Presenting to Hospital with Diarrhea from Lambaréné, Gabon. Am J Trop Med Hyg. 2021;105: 254–260. doi:10.4269/ajtmh.20-1290

7.	Moyo SJ, Kommedal Ø, Blomberg B, Hanevik K, Tellevik MG, Maselle SY, et al. Comprehensive Analysis of Prevalence, Epidemiologic Characteristics, and Clinical Characteristics of Monoinfection and Coinfection in Diarrheal Diseases in Children in Tanzania. American Journal of Epidemiology. 2017;186: 1074–1083. doi:10.1093/aje/kwx173

8.	Debes AK, Xiao S, Liu J, Shaffer A, Scalzo P, Guenou E, et al. Characterization of Enteric Disease in Children by Use of a Low-Cost Specimen Preservation Method. J Clin Microbiol. 2021;59: e0170321. doi:10.1128/JCM.01703-21

9.	Page NA, Seheri LM, Groome MJ, Moyes J, Walaza S, Mphahlele J, et al. Temporal association of rotavirus vaccination and genotype circulation in South Africa: Observations from 2002 to 2014. Vaccine. 2018;36: 7231–7237. doi:10.1016/j.vaccine.2017.10.062

10.	Kotloff KL, Blackwelder WC, Nasrin D, Nataro JP, Farag TH, van Eijk A, et al. The Global Enteric Multicenter Study (GEMS) of diarrheal disease in infants and young children in developing countries: epidemiologic and clinical methods of the case/control study. 2012;55 Suppl 4: S232-45. doi:10.1093/cid/cis753

11.	Platts-Mills JA, Amour C, Gratz J, Nshama R, Walongo T, Mujaga B, et al. Impact of Rotavirus Vaccine Introduction and Postintroduction Etiology of Diarrhea Requiring Hospital Admission in Haydom, Tanzania, a Rural African Setting. Clinical Infectious Diseases. 2017;65: 1144–1151. doi:10.1093/cid/cix494

12.	Ashorn P, Alho L, Ashorn U, Cheung YB, Dewey KG, Gondwe A, et al. Supplementation of Maternal Diets during Pregnancy and for 6 Months Postpartum and Infant Diets Thereafter with Small-Quantity Lipid-Based Nutrient Supplements Does Not Promote Child Growth by 18 Months of Age in Rural Malawi: A Randomized Controlled Trial. The Journal of Nutrition. 2015;145: 1345–1353. doi:10.3945/jn.114.207225

13.	Lambrecht NJ, Wilson ML, Bridges D, Eisenberg JNS, Adu B, Baylin A, et al. Ruminant-Related Risk Factors are Associated with Shiga Toxin–Producing Escherichia coli Infection in Children in Southern Ghana. The American Journal of Tropical Medicine and Hygiene. 2022;106: 513–522. doi:10.4269/ajtmh.21-0550

14.	MAL-ED Network Investigators. The MAL-ED study: a multinational and multidisciplinary approach to understand the relationship between enteric pathogens, malnutrition, gut physiology, physical growth, cognitive development, and immune responses in infants and children up to 2 years of. Clinical infectious diseases. 2014;59 Suppl 4: S193-206. doi:10.1093/cid/ciu653

15.	Bill & Melinda Gates Foundation. Improved Biomarkers for the Assessment of Environmental Enteropathy. In: Global Grand Challenges [Internet]. 2012 [cited 18 Nov 2020]. Available: https://gcgh.grandchallenges.org/grant/improved-biomarkers-assessment-environmental-enteropathy

16.	Chisenga CC, Bosomprah S, Makabilo Laban N, Mwila- Kazimbaya K, Mwaba J, Simuyandi M, et al. Aetiology of Diarrhoea in Children Under Five in Zambia Detected Using Luminex xTAG Gastrointestinal Pathogen Panel. Pediatric Infectious Diseases: Open Access Scitechnol Biosoft Pvt. Ltd.; 2018. doi:10.21767/2573-0282.100064

17.	Pelkonen T, Dos Santos MD, Roine I, Dos Anjos E, Freitas C, Peltola H, et al. Potential Diarrheal Pathogens Common Also in Healthy Children in Angola. Pediatric Infectious Disease Journal. 2018;37: 424–428. doi:10.1097/INF.0000000000001781

18.	Lima AAM, Oliveira DB, Quetz JS, Havt A, Prata MMG, Lima IFN, et al. Etiology and severity of diarrheal diseases in infants at the semiarid region of Brazil: A case-control study. Gurley E, editor. PLOS Neglected Tropical Diseases. 2019;13: e0007154. doi:10.1371/journal.pntd.0007154

19.	Bhandari N, Rongsen-Chandola T, Bavdekar A, John J, Antony K, Taneja S, et al. Efficacy of a monovalent human-bovine (116E) rotavirus vaccine in Indian infants: A randomised, double-blind, placebo-controlled trial. The Lancet. 2014;383: 2136–2143. doi:10.1016/S0140-6736(13)62630-6

20.	Humphrey JH, Mbuya MNNN, Ntozini R, Moulton LH, Stoltzfus RJ, Tavengwa N V., et al. Independent and combined effects of improved water, sanitation, and hygiene, and improved complementary feeding, on child stunting and anaemia in rural Zimbabwe: a cluster-randomised trial. The Lancet Global Health. 2019;7: e132–e147. doi:10.1016/S2214-109X(18)30374-7

21.	Okada K, Wongboot W, Kamjumphol W, Suebwongsa N, Wangroongsarb P, Kluabwang P, et al. Etiologic features of diarrheagenic microbes in stool specimens from patients with acute diarrhea in Thailand. Scientific Reports. 2020;10: 4009. doi:10.1038/s41598-020-60711-1

22.	Gaensbauer JT, Lamb M, Calvimontes DM, Asturias EJ, Kamidani S, Contreras-Roldan IL, et al. Identification of Enteropathogens by Multiplex PCR among Rural and Urban Guatemalan Children with Acute Diarrhea. The American journal of tropical medicine and hygiene. 2019;101: 534–540. doi:10.4269/ajtmh.18-0962

23.	Chard AN, Baker KK, Tsai K, Levy K, Sistrunk JR, Chang HH, et al. Associations between soil-transmitted helminthiasis and viral, bacterial, and protozoal enteroinfections: a cross-sectional study in rural Laos. Parasites & Vectors. 2019;12: 216. doi:10.1186/s13071-019-3471-2

24.	ICF International. Demographic and Health Surveys (various, 2000-2021). Rockville, Maryland, USA: ICF International; 2021.

25.	UNICEF. Multiple Indicator Cluster Surveys (various, 2000-2021). New York, NY: UNICEF; 2021.

26.	World Health Organization. WHO Anthro Survey Analyser and other tools. In: Child Growth Standards [Internet]. 6 Mar 2019 [cited 10 Dec 2021]. Available: https://www.who.int/tools/child-growth-standards/software

27.	Bhattacharjee N V., Schaeffer LE, Marczak LB, Ross JM, Swartz SJ, Albright J, et al. Mapping exclusive breastfeeding in Africa between 2000 and 2017. Nature Medicine. 2019;25. doi:10.1038/s41591-019-0525-0

28.	Kinyoki DK, Osgood-Zimmerman AE, Pickering B V., Schaeffer LE, Marczak LB, Lazzar-Atwood A, et al. Mapping child growth failure across low- and middle-income countries. Nature. 2020;577: 231–234. doi:10.1038/s41586-019-1878-8

29.	Florey L, Taylor C. Using household survey data to explore the effects of improved housing conditions on malaria infection in children in Sub-Saharan Africa. Rockville, Maryland, USA: ICF International; 2016 Aug.

30.	Knee J, Sumner T, Adriano Z, Berendes D, de Bruijn E, Schmidt W-P, et al. Risk factors for childhood enteric infection in urban Maputo, Mozambique: A cross-sectional study. Mejia R, editor. PLOS Neglected Tropical Diseases. 2018;12: e0006956. doi:10.1371/journal.pntd.0006956

31.	Graetz N, Woyczynski L, Wilson KF, Hall JB, Abate KH, Abd-Allah F, et al. Mapping disparities in education across low- and middle-income countries. Nature. 2019. doi:10.1038/s41586-019-1872-1

32.	World Health Organization, UNICEF. Joint Monitoring Programme (JMP) for Water Supply and Sanitation. 2021 [cited 4 Aug 2021]. Available: https://washdata.org/

33.	Deshpande A, Miller-Petrie MK, Lindstedt PA, Baumann MM, Johnson KB, Blacker BF, et al. Mapping geographical inequalities in access to drinking water and sanitation facilities in low-income and middle-income countries, 2000–17. The Lancet Global Health. 2020;8: e1162–e1185. doi:10.1016/S2214-109X(20)30278-3

34.	Weiss DJ, Nelson A, Gibson HS, Temperley W, Peedell S, Lieber A, et al. A global map of travel time to cities to assess inequalities in accessibility in 2015. Nature. 2018;553: 333–336. doi:10.1038/nature25181

35.	Ramankutty N, Evan AT, Monfreda C, Foley JA. Farming the planet: 1. Geographic distribution of global agricultural lands in the year 2000. Global Biogeochemical Cycles. 2008;22: n/a-n/a. doi:10.1029/2007GB002952

36.	Natural Earth. Rivers and Lakes Centerlines 4.1.0. 2021.

37.	Hastings DA, Dunbar PK. Global Land One-kilometer Base Elevation (GLOBE) Digital Elevation Model, Documentation, Volume 1.0. Boulder, Colorado; 1999.

38.	U.S. Geological Survey. Landsat Enhanced Vegetation Index. In: Landsat Missions [Internet]. 2021 [cited 14 Dec 2021]. Available: https://www.usgs.gov/landsat-missions/landsat-enhanced-vegetation-index

39.	The Food and Agriculture Organization (FAO), International Institute of Applied Systems Analysis. Global Agro-ecological Zones (GAEZ v3.0). Rome, Italy and Laxenburg, Austria: FAO & IIASA; 2012.

40.	Wildlife Conservation Society, Center for International Earth Science Information Network - CIESIN. Last of the Wild Project, Version 2, 2005 (LWP-2): Global Human Footprint Dataset (Geographic). Palisades, NY: NASA Socioeconomic Data and Applications Center (SEDAC); 2005.

41.	Siebert S, Döll P, Hoogeveen J, Faures J-M, Frenken K, Feick S. Development and validation of the global map of irrigation areas. Hydrology and Earth System Sciences. 2005;9: 535–547. doi:10.5194/hess-9-535-2005

42.	Tatem AJ. WorldPop, open data for spatial demography. Scientific Data. 2017;4: 170004. doi:10.1038/sdata.2017.4

43.	Pesaresi M, Ehrlich D, Stefano F, Florcyk A, Freire SMC, Halkia S, et al. Operating procedure for the production of the Global Human Settlement Layer from Landsat data of the epochs 1975, 1990, 2000, and 2014 | EU Science Hub. 2016.

44.	Rodell M, Houser PR, Jambor U, Gottschalck J, Mitchell K, Meng C-J, et al. The Global Land Data Assimilation System. http://dx.doi.org/101175/BAMS-85-3-381. 2004.

45.	ESRI. ArcGIS Desktop: Release 10.8. Redlands, CA: Environmental Systems Research Institute; 2019. Available: https://desktop.arcgis.com/en/desktop/

46.	Colston JM, Zaitchik B, Kang G, Peñataro Yori P, Ahmed T, Lima A, et al. Use of earth observation-derived hydrometeorological variables to model and predict rotavirus infection (MAL-ED): a multisite cohort study. The Lancet Planetary Health. 2019;3: S2542-5196. doi:10.1016/S2542-5196(19)30084-1

47.	Bhandari D, Bi P, Dhimal M, Sherchand JB, Hanson-Easey S. Non-linear effect of temperature variation on childhood rotavirus infection: A time series study from Kathmandu, Nepal. Science of the Total Environment. 2020;748. doi:10.1016/j.scitotenv.2020.141376

48.	Aslam A, Gossman WG. Shigella (Shigellosis). StatPearls. StatPearls Publishing; 2018.

49.	Statistics Botswana, Ministry of Finance and Development Planning. Botswana Family Health Survey 2007 - 08. Botswana; 2008. Report No.: bwa-sb-bfhs-2007-2008-v1. Available: https://catalog.ihsn.org/index.php/catalog/7414

50.	Ministério da Saúde, Brasil. Pesquisa Nacional de Demografia e Saúde da Criança e da Mulher (PNDS). Brasília, DF: Ministério da Saúde, Brasil; 2006. Available: https://bvsms.saude.gov.br/bvs/pnds/

51.	The World Bank. The world by region. In: SDG Atlas 2017 [Internet]. 2017 [cited 10 Dec 2021]. Available: https://datatopics.worldbank.org/sdgatlas/archive/2017/the-world-by-region.html

52.	Colston JM. Seasonality and Hydrometeorological Predictors of Rotavirus Infection in an Eight-Site Birth Cohort Study: Implications for Modeling and Predicting Pathogen-Specific Enteric Disease Burden. Johns Hopkins University. 2018. Available: http://jhir.library.jhu.edu/handle/1774.2/61085

53.	Stevens GA, Alkema L, Black RE, Boerma JT, Collins GS, Ezzati M, et al. Guidelines for Accurate and Transparent Health Estimates Reporting: the GATHER statement. The Lancet. 2016;388: e19–e23. doi:10.1016/S0140-6736(16)30388-9

54.	Stewart LA, Clarke M, Rovers M, Riley RD, Simmonds M, Stewart G, et al. Preferred Reporting Items for Systematic Review and Meta-Analyses of individual participant data: the PRISMA-IPD Statement. JAMA. 2015;313: 1657–1665. doi:10.1001/jama.2015.3656

1. These two binary variables were combined into a categorical variable with four values and asymptomatic-community surveillance as the reference category.
